# Prioritizing allocation of COVID-19 vaccines based on social contacts increases vaccination effectiveness

**DOI:** 10.1101/2021.02.04.21251012

**Authors:** Jiangzhuo Chen, Stefan Hoops, Achla Marathe, Henning Mortveit, Bryan Lewis, Srinivasan Venkatramanan, Arash Haddadan, Parantapa Bhattacharya, Abhijin Adiga, Anil Vullikanti, Aravind Srinivasan, Mandy L Wilson, Gal Ehrlich, Maier Fenster, Stephen Eubank, Christopher Barrett, Madhav Marathe

## Abstract

We study allocation of COVID-19 vaccines to individuals based on the structural properties of their underlying social contact network. Even optimistic estimates suggest that most countries will likely take 6 to 24 months to vaccinate their citizens. These time estimates and the emergence of new viral strains urge us to find quick and effective ways to allocate the vaccines and contain the pandemic. While current approaches use combinations of age-based and occupation-based prioritizations, our strategy marks a departure from such largely aggregate vaccine allocation strategies. We propose a novel agent-based modeling approach motivated by recent advances in (*i*) science of real-world networks that point to efficacy of certain vaccination strategies and (*ii*) digital technologies that improve our ability to estimate some of these structural properties. Using a realistic representation of a social contact network for the Commonwealth of Virginia, combined with accurate surveillance data on spatio-temporal cases and currently accepted models of within- and between-host disease dynamics, we study how a limited number of vaccine doses can be strategically distributed to individuals to reduce the overall burden of the pandemic. We show that allocation of vaccines based on individuals’ degree (number of social contacts) and total social proximity time is *significantly more effective* than the currently used age-based allocation strategy in terms of number of infections, hospitalizations and deaths. Our results suggest that in just two months, by March 31, 2021, compared to age-based allocation, the proposed degree-based strategy can result in *reducing an additional 56–110k infections, 3.2–5.4k hospitalizations, and 700–900 deaths just in the Commonwealth of Virginia. Extrapolating these results for the entire US, this strategy can lead to 3–6 million fewer infections, 181–306k fewer hospitalizations, and 51–62k fewer deaths compared to age-based allocation.* The overall strategy is robust even: (*i*) if the social contacts are not estimated correctly; (*ii*) if the vaccine efficacy is lower than expected or only a single dose is given; (*iii*) if there is a delay in vaccine production and deployment; and (*iv*) whether or not non-pharmaceutical interventions continue as vaccines are deployed. For reasons of implementability, we have used degree, which is a simple structural measure and can be easily estimated using several methods, including the digital technology available today. These results are significant, especially for resource-poor countries, where vaccines are less available, have lower efficacy, and are more slowly distributed.

## 1 Introduction

New vaccines typically take a decade to develop and distribute, but vaccines for COVID-19, the disease caused by the novel coronavirus SARS-CoV-2, have been developed in record time to help mitigate the raging pandemic. As of February 13, 2021, the reported number of confirmed cases and deaths in the US stand at 27M and 467K; the reported number of confirmed cases and deaths worldwide stand at 103M and 2.3M respectively.^1^. These numbers are likely to go up substantially in the coming months. Vaccines offer a safe and effective way to contain the pandemic quickly. However, the supply of COVID-19 vaccines is limited, so the challenge now is the distribution of these vaccines in a timely manner to bring the pandemic under control. If we have a sufficient number of vaccines to immunize 70-90% of the people in the United States (US), protection can be offered to both individuals who are immunized and those who are unimmunized through herd immunity.

In the next 3 months, the US is expected to have a total of only 100 million vaccines, which is sufficient to immunize only 30% of the population (15% if we account for two doses) and thus cannot provide herd immunity. Lacking that, the current focus of the vaccination is to protect individuals who are at a high risk of infection and mortality, as well as critical workers.

Vaccination priority is complex and intertwined with age, race, occupation, health equity, geography, and politics. Data shows that COVID-19 disproportionately affects older adults, Blacks, Hispanics, American Indians, gig and wage workers, and individuals with comorbidities. Many of these attributes are also correlated with low socio-economic status (SES), and high social vulnerability. There can be many criteria for prioritization, for example: (*i*) risk of infection; (*ii*) risk of death; (*iii*) risk of transmission if infected; and (*iv*) occupation, such as healthcare workers, teachers, cashiers, etc. Estimating the consequences of different prioritization strategies is further complicated by production limitations, requiring a vaccine schedule to be specified for each. Additionally, vaccine distribution requires complex logistical support, such as cold-chain storage, transportation, qualified personnel, and scheduling etc. for any prioritization scheme to achieve its results in an effective and equitable manner. See [4] for a comprehensive discussion on this topic. The US Centers for Disease Control and Prevention (CDC) has announced a prioritization order based on the Advisory Committee on Immunization Practices (ACIP). It recommends healthcare personnel and long-term facility care residents be vaccinated first; followed by frontline essential workers, and those aged 75 years and older because they are at a higher risk of hospitalization, illness and death; followed by those aged 65-74 years; followed by those aged 16-64 years with underlying medical conditions and other essential workers. Although most states in the US follow a similar phased approach, there are subtle, but important, variations on who gets vaccinated first.

### Our contributions

The current rate of vaccination in the US and other countries suggests that it can take between 6-24 months to complete vaccination campaigns for much of the world. At the same time, the discovery of multiple variant strains implies a rapid acceleration of the pandemic in several parts of the world. The number of new strains are likely to increase with increased prevalence. Thus a natural question to study is the following: *can we prioritize vaccine distribution so as to significantly reduce the overall burden of COVID-19 quickly?*

We propose prioritization schemes based on properties of individuals within social contact networks with the goal of bending the pandemic curve and improving overall pandemic outcome. We synthesize a digital twin of Virginia, which is a detailed social contact network model for the Commonwealth of Virginia (8 million individuals), and use an agent-based model (ABM) to study the effectiveness of various prioritization schemes. In contrast to other such networks, our networks incorporate detailed information about the population, their activities and the built infrastructure. Further information on how such a digital twin is constructed and its structural properties can be found in Section A.1. The ABM simulates disease propagation and a complex set of interventions, including various non-pharmaceutical interventions and vaccine allocation schemes.

Our prioritization schemes based on simple, individual-based yet computable, structural properties of the underlying social contact network are motivated by: (*i*) recent advances in network science that have studied such schemes in more abstract settings; (*ii*) our ability to construct detailed, realistic social contact networks at scale; (*iii*) our ability to simulate and assess such strategies even for complex disease transmission models and public health control measures; and (*iv*) recent progress in development of digital apps that can be used for measuring structural properties in large populations relatively accurately, rendering such schemes potentially operationalizable.

Our prioritization schemes can be stated simply as follows: *vaccinate individuals who typically exhibit high social contact (degree or total contact time in the social contact network)*. Some key points to note: (*i*) we focus on simple network structural properties that can be estimated in a privacy-preserving way, (*ii*) we do not insist on strict ordering of individuals nor an exact estimation of their social contacts, and (*iii*) while our *analysis* uses a realistic representation of the social contact networks, implementation of the policy does not require one to *synthesize* the social network.

There is folklore that degree based heuristics to allocate vaccines often work well. The folklore is based on mathematical results for highly structured random networks or on computational experiments based on relatively simple class of social contact networks [10,21,46,56]. But the folklore has never been tested in time-varying realistic social contact networks such as the one constructed here and intended to capture the network evolution due to adaptive NPIs and vaccine allocation that is undertaken in a time varying manner. Our results show for the first time that degree based heuristics are likely to work even for such time-varying social contact networks; see Sections H and E for further discussions on this topic.

Our results suggest that in just two months (i.e. by the end of March 2021), compared to age-based allocation, the proposed degree-based strategy can result in averting an *additional 56– 110k infections (8–16%), 3.2–5.4k hospitalizations (8–13%), and 700–900 deaths (6–8%) just in the state of Virginia. Extrapolating these results per capita for the entire US, we estimate this strategy will lead to 3–6 million fewer infections, 181–306k fewer hospitalizations, and 51–62k fewer deaths compared to the age-based allocation.* The results continue to hold qualitatively and show that we can avert many more infections, hospitalizations, and deaths even if the current social distancing measures are relaxed. Furthermore, similar results hold even for vaccines with 50% efficacy; this is important, as most resource-poor countries do not have access to high efficacy vaccines at this point in time. The basic intuition behind our results is that vaccinating individuals with high degree not only protects them but also confers significant protection to individuals who come in close proximity in their contact network.

A natural question is: *how can such individuals be identified?* This might be done by objectively determining who they are and seeking them out. Alternatively, at a time of interview, a person could be designated as “high degree” through identifying data or proxy characteristics to necessary statistical precision that show the individual belongs to such a critical group identified by the model. We discuss how currently deployed digital contact tracing apps can be modified in a very simple manner to achieve the goal of identifying high degree individuals (Section E). Such individuals can also be identified by observing that certain occupations naturally lead to a high level of social interactions. Our methods are robust to partial mis-estimation of these social contacts and their implementation does *not* require access to the social contact network.

## 2 Experiment Settings and Design

For the experiments, we use an agent-based simulation model, EpiHiper, which is described in Appendix F and has been used in previous studies [18]. The simulation’s input parameters specify the population demographics and contact network, COVID-19 disease model, initial configuration *S*_0_, non-pharmaceutical interventions (NPIs), and vaccination schedule. The simulation output is a dendrogram: a directed graph that tells us who infects whom and on what day. From the output data, we can compute many epidemiological measures such as daily new infections, cumulative infections, prevalence in each age group, total hospitalizations, and deaths, as well as many other measures.

### 2.1 Simulation parameterization

These studies use a synthetic population and contact network for Virginia, which is described in Appendix A.1. The initial conditions are calibrated to the conditions in Virginia as of January 1, 2021. Every simulation is run for 90 days, until March 31, 2021. Since the simulations are stochastic, each simulation is repeated for 30 replicates, and distributions of the measures are computed. The boxplots and curves in figures presented in Section 3 (Figure 1 through 11) are all based on data from 30 replicates. The curves show an uncertainty of one standard deviation above and below the mean.

**Figure 1:**
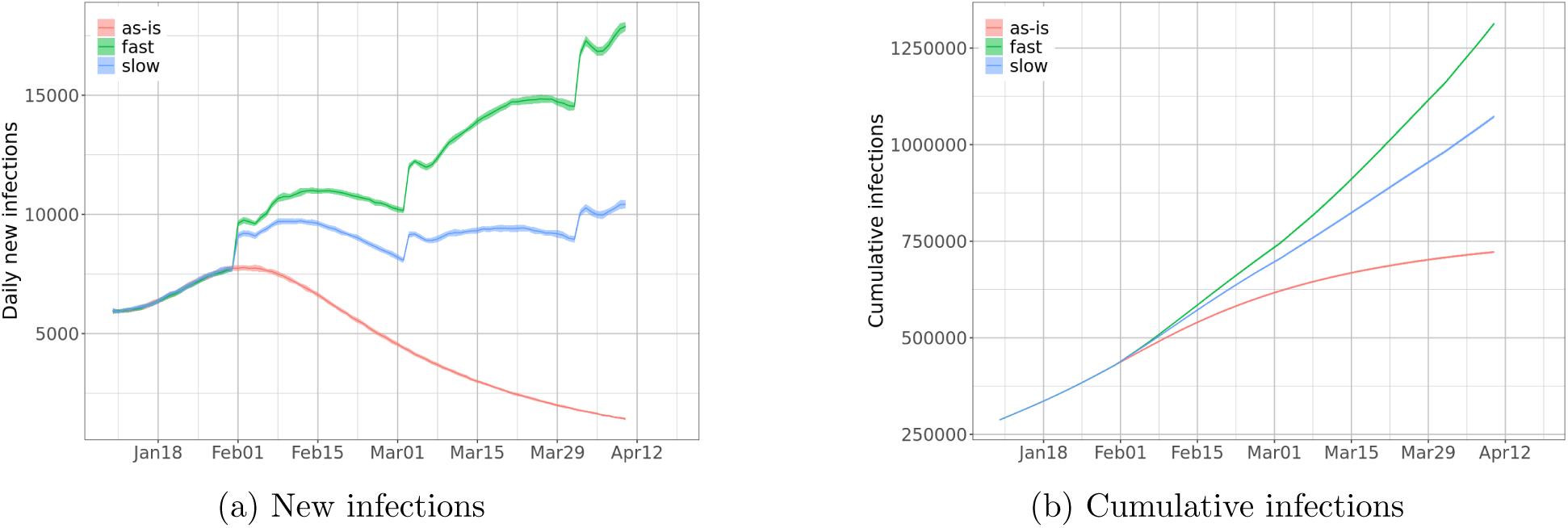
Incidence without vaccination under different NPI relaxation scenarios: (a) daily number of new infections; (b) cumulative number of infections. The sudden surge in new infections in the beginning of each month is caused by NPI relaxation. Without NPI relaxation, the incidence reaches peak in early February then starts decreasing. With slow relaxation, daily incidence increases then fluctuates. With fast relaxation, the incidence keeps rising.

#### Disease model

The disease model is the *best guess version* of “COVID-19 Pandemic Planning Scenarios” prepared by the US Centers for Disease Control and Prevention (CDC) SARS-CoV-2 Modeling Team [16] and has been used by multiple researchers in their papers. It is an SEIR model where state transitions follow the parameters as defined in the document. The disease states and transition paths are shown in Figure 24. Individuals of different age groups have different infectivity and susceptibility; dwell time distributions and state transition probability distributions are stratified by the following age groups: preschool (0-4 years), students (5-17), adults (18-49), older adults (50-64) and seniors (65+). Furthermore, individuals that are vaccinated have different disease parameter values than those that are not vaccinated. Detailed parameterization for unvaccinated individuals is summarized in Appendix G.

#### Initializations

The simulations are initialized at the county level by age group using the detailed data of confirmed cases from [60]. The initialization specifies the health state of each individual. Based on county-level cumulative confirmed cases through December 19, 2020, we derive the number of prior infections in each county by scaling the cumulative number by a case ascertainment ratio of 3 (i.e., only one third of all infections are reported), then computing the number of prior infections in each age group of this county using the age distribution in cases. We randomly choose individuals in each age group in each county and set their health states to `recovered` to reflect that they have already been infected. Based on county-level daily confirmed cases from December 20, 2020 to January 5, 2021, we derive the number of individuals that are infected each day by the same scaling, and seed the simulation by setting randomly chosen individuals to *exposed* by day in each age group of each county.

#### Non-pharmaceutical interventions

We consider four NPIs: (*i*) *Infectivity reduction (IR)*. Infectivity is universally reduced (by 60%) through preventive behavior, e.g., mask wearing and hand washing. (*ii*) *Generic social distancing (GSD)*. A fraction (25%) of the population chooses to reduce non-essential (shopping, religion, and other) activities. (*iii*) *Virtual learning (VL)*. A fraction (50%) of K-12 students choose virtual learning. (*iv*) *Voluntary home isolation of symptomatic cases (VHI)*. With probability 75%, a symptomatic person chooses to stay home for 14 days, reducing the weights on household contacts by 50%. For this person, all outside contacts are disabled and at-home contacts are reduced by 50% temporarily during these 14 days.

#### Scenarios based on relaxing social distancing measures

We assume that these NPIs are in place when a simulation starts, but adherence may change during the simulation. We consider three scenarios for adherence to the NPIs:

- **As-is.** NPI parameters remain the same for the duration of the simulation.
- **Slow relaxation.** NPI parameters change every 30 days from January 30, 2021, so that in 7 months, infectivity reduction decreases from 60% to 10%, generic social distancing decreases from 25% to 10%, and virtual learning decreases from 50% to 25%. Note that this is used to specify the speed of relaxation. Nevertheless the results are only reported for the period until end of March.
- **Fast relaxation.** NPI parameters change every 30 days from January 30, 2021, so that in 5 months, they reach the same levels as in the *slow relaxation* scenario.

### 2.2 Vaccination: supply, schedule and priority groups

#### Vaccine schedule

As of this writing, we expect 400 million doses to be delivered by Pfizer-BioNTech and Moderna to the US by the end of July 2021, enough to vaccinate 200 million people. By assuming that 25 million people can be vaccinated per month, starting from late December 2020 until late July 2021, and that vaccines are allocated to all states proportional to population size, we consider a vaccination schedule as shown in Table 1, where 650K people are vaccinated per month in Virginia, and a schedule where they are vaccinated at half this rate. Therefore we consider three vaccination schedules: *none* (no vaccination), *fast* (vaccinating 650K people per month), and *slow* (vaccinating 325K people per month). The later schedule is intended to capture the current challenges faced in distributing the vaccines to individuals. For simplicity, all individuals vaccinated during each month are assumed to be vaccinated on the first day of that month; spreading the vaccines over the month does not change the overall results by much.

**Table 1:** Cumulative number of individuals vaccinated in each month of 2021. Note that in our experiment, where simulations run until the end of March, we consider vaccinations up to March only.

| vaccination up to | US (million) | Virginia (thousand) |
| --- | --- | --- |
| Jan | 25 | 650 |
| Feb | 50 | 1300 |
| Mar | 75 | 1950 |
| Apr | 100 | 2600 |
| May | 125 | 3250 |
| Jun | 150 | 3900 |
| Jul | 175 | 4550 |
| Aug | 200 | 5200 |

#### Vaccine efficacy

Overall vaccine efficacy is characterized by three numbers: (*i*) *e_I_*, efficacy against infection; (*ii*) *e_D_*, efficacy against severe illness (requiring hospitalization or leading to death) given infection; and (*iii*) *e_T_*, efficacy against onward transmission given infection. We assume that *e_I_* = 90% and *e_D_* = 50% starting only 21 days after vaccination. In our sensitivity analyses, we also consider *e_I_* = 50%. In all cases, we ignore *e_T_*.

#### Vaccination prioritization

The Pfizer-BioNTech vaccine and the Moderna vaccine are recommended for people aged at least 16 years and at least 18 years, respectively. In the experiments, we only allocate vaccines to people who are at least 18 years old. Among those people, we consider the following prioritization strategies.

- *No priority*. Everyone 18+ years old is vaccinated with the same probability. This is our baseline strategy.
- *Essential workers*. This strategy targets those who work for medical, care facilitation, retail, education, military, and government.
- *Older people*. This strategy prioritizes those who are at least 50 years old.
- *High degree*. Degree of an individual is the number of contacts per day. This strategy targets those in the top quartile among all 18+ years old in terms of degree.
- *Long total contact (also denoted as weighted degree)*. Weighted degree of an individual is the total contact time this individual has with other people in a day. This strategy targets those in the top quartile among all 18+ years old in terms of weighted degree.

Most vaccines are allocated to the targeted groups, but we allow some to be given to other groups. This accounts for potential inaccuracy and precision in identifying and locating the targeted people. For example, since we do not know people’s daily number of contacts, which may vary, we can only estimate it using proxy attributes, such as age, household size and occupation, or from data collected through digital devices. We consider the following rates of enforcement: 100%, 80%, and 60%.

### 2.3 Experimental design

The design consists of 4 factors: (*i*) 3 adherence scenarios (as-is, slow relaxation, fast relaxation); (*ii*) 3 vaccination schedules (none, fast, slow); (*iii*) 5 prioritization targets (no priority, essential workers, older people, high degree, high weighted degree); and (*iv*) 3 levels of priority enforcement (100%, 80%, 60%). Combining (*iii*) and (*iv*) we have the baseline (no-priority) plus 12 prioritized strategies named according to the target group (essential, old-age, high degree, high weighted degree) and the fraction of vaccine given to the target group (100%, 80%, 60%), e.g., “essential 100%” or “high degree 60%”. We also consider vaccines with a 50% efficacy against infection (*e_I_*) and compare the effectiveness of degree-based vaccinations under this assumption against that under 90% efficacy.

## 3 Results and Analysis

In Figure 1, we show daily new infections under three scenarios (as-is, slow relaxation and fast relaxation) *without vaccination*. If NPI adherence can be maintained, then we expect infections to decrease after January. With slow relaxation, the infections will fluctuate around a level that will be a little higher than the current level. With fast relaxation, the infections show a steady increase in the next three months. The sharp increase every 30 days is caused by the implementation of the relaxation of NPIs and does not have any influence on the results presented.

### 3.1 Effectiveness of degree- and weighted degree-based strategies

Prioritizing vaccinations based on individual degree and weighted degree are extremely effective in controlling the pandemic. In particular, depending on the scenario, the reductions in the number of infections and hospitalizations by these schemes are over 50% more than the reductions from the age-based prioritization schemes. For example, assuming that the current non-pharmaceutical interventions remain at the same level over the next few months, our experiment shows that by the end of March 2021, degree-based schemes can result in 56–110k fewer infections, 3.2–5.4k fewer hospitalizations, and 700–900 fewer deaths in the state of Virginia, compared to age-based schemes. Note that the ranges come from different levels of priority enforcement (three levels for both age-based and degree-based schemes). Figure 2 shows the estimated reductions by one of the age-based schemes and the further reductions by one of the degree-based schemes. Extrapolating these results for the entire US, we estimate that degree-based schemes will lead to 3–6 million fewer infections, 181–306k fewer hospitalizations, and 51–62k fewer deaths by the end of March, compared to age-based schemes. If the NPIs are relaxed, the reductions in infections, hospitalizations, and mortality are even more substantial. This implies that when conditions worsen, the marginal gains from a more effective strategy are even higher.

**Figure 2:**
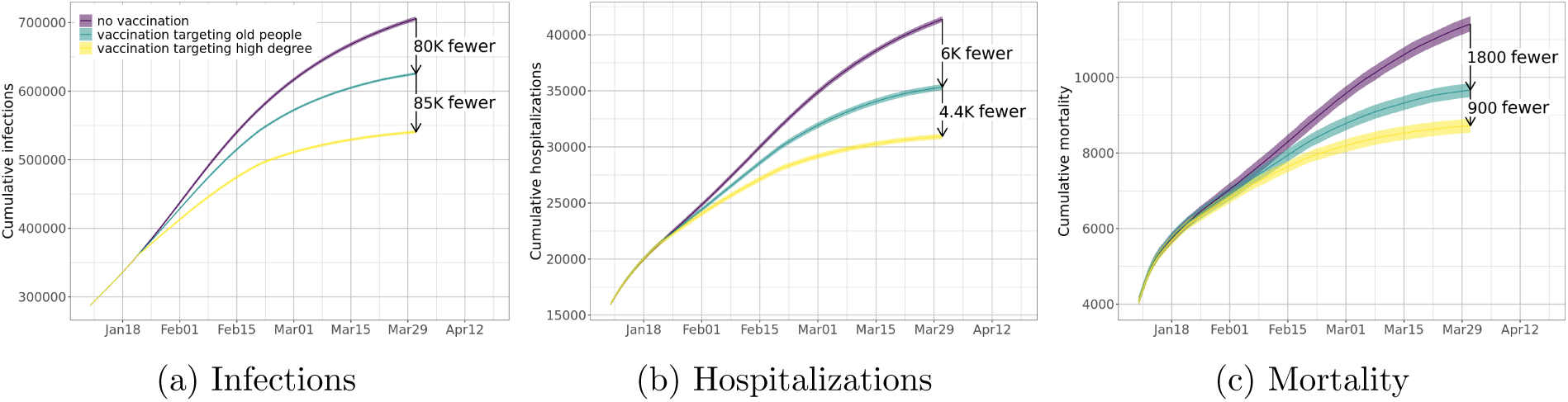
Vaccination targeting old people can reduce (a) total infections, (b) total hospitalizations, and (c) total mortality significantly, assuming current non-pharmaceutical interventions remain at the same level. Vaccination targeting high degree people can further reduce total infections, hospitalizations, and mortality. Numbers in the plots show total reductions up to the end of March 2021. Note that the “no vaccination” curves in (a) is the same as the “as-is” curve in Figure 1b.

Figure 3 compares incidence reduction up to March 31, 2021, under different prioritization strategies for the vaccine distribution schedule given in Table 1, also known as the *fast* schedule. We find that all strategies targeting either essential workers or high degree people outperform the no-priority distribution. The degree-based strategies reduce incidence more than any other strategy. For example, with no NPI relaxation (as-is), all degree-based strategies can reduce infections by over 20% while all other strategies can reduce infections by at most 20%. Strategies targeting older people perform worse than the no-priority distribution in terms of reducing incidence. Similar results are obtained for the *slow* vaccine distribution schedule, as shown in Figure 4. One reason to consider weighted degree-based heuristics is that they are potentially easier to implement in the current digital apps, we will discuss this further in later sections. **All degree-based strategies outperform the other strategies.**

**Figure 3:**
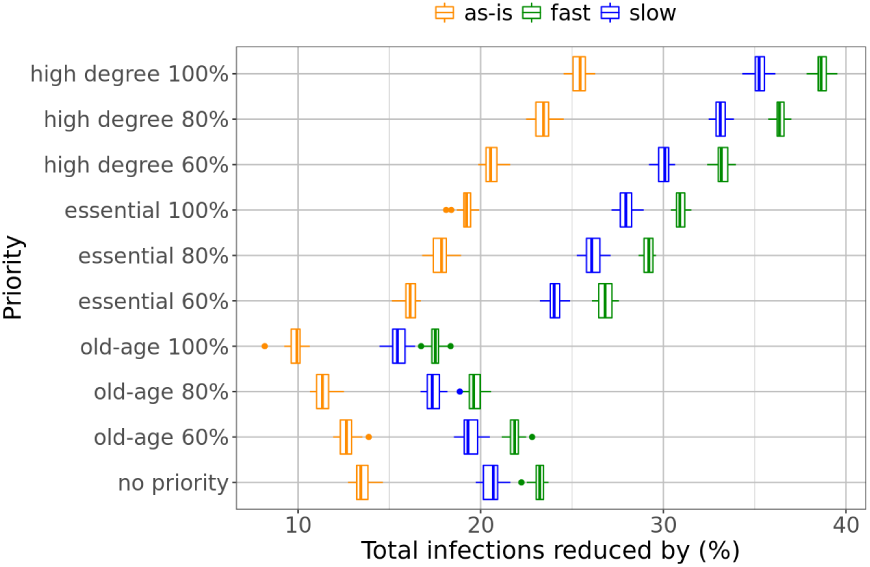
Total reduction in incidence under the fast vaccine distribution schedule. Degree-based strategies outperforms all other ones, while age-based strategies are outperformed by all other ones.

**Figure 4:**
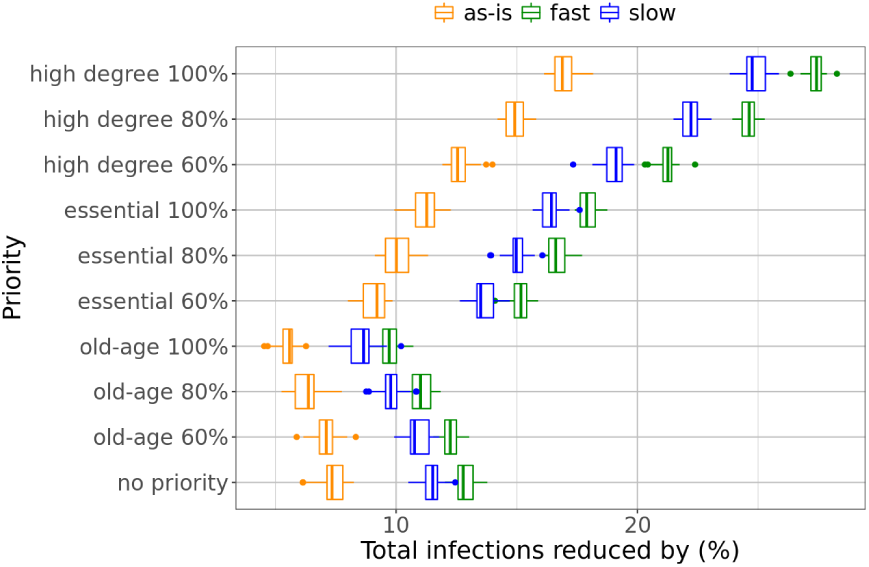
Total reduction in incidence under the slow vaccine distribution schedule. Degree-based strategies still reduce more infections than other strategies and are more effective if accuracy is higher.

Targeting high degree people is also the most effective strategy for reducing mortality. Prioritization of older people is effective in reducing mortality compared to other strategies, but not when compared to a high degree strategy. This is shown in Figures 5 and 6. Figure 7 shows that prioritizing people with high weighted degree (total contact durations) is even more effective than prioritizing those with high degree. For example, Figure 7a shows that, with no relaxation of NPIs, targeting people of high weighted degree can reduce infections by about 23-30%, compared to targeting high degree people, which can reduce infections by about 21-26%. In the case where NPIs are relaxed, the strategy prioritizing high weighted degree can cause over 40% reduction in infections if it can be implemented with high precision.

**Figure 5:**
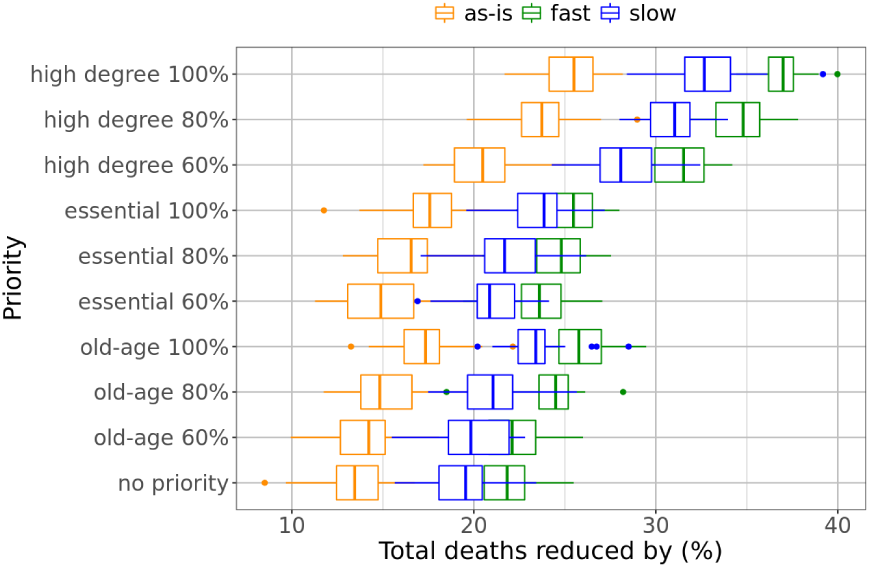
Total reduction in mortality under the fast vaccine distribution schedule. While degree-based strategies continue to perform better than other ones in reducing mortality, age-based strategies seem to be more effective than the baseline.

**Figure 6:**
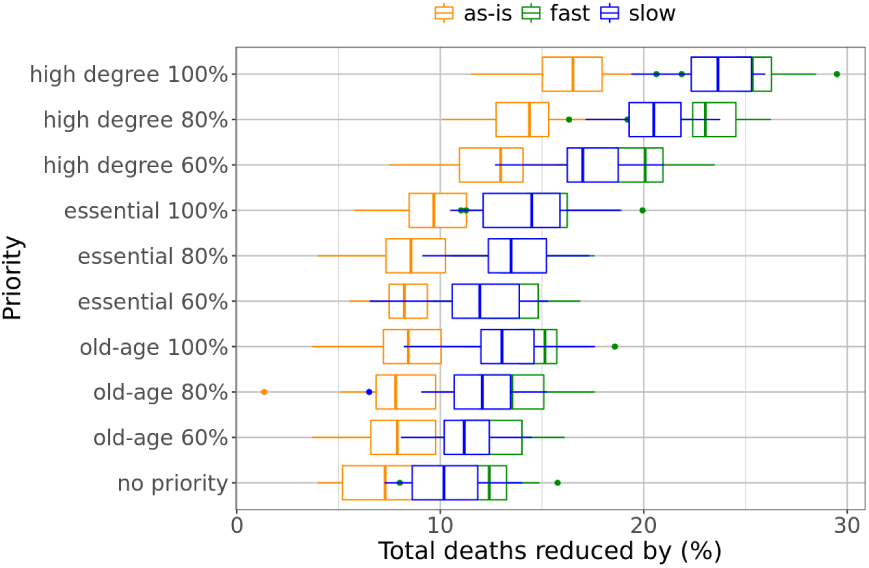
Total reduction in mortality under the slow vaccine distribution schedule. Observations in the fast vaccine distribution case remain true, except that the reductions now have smaller sizes.

**Figure 7:**
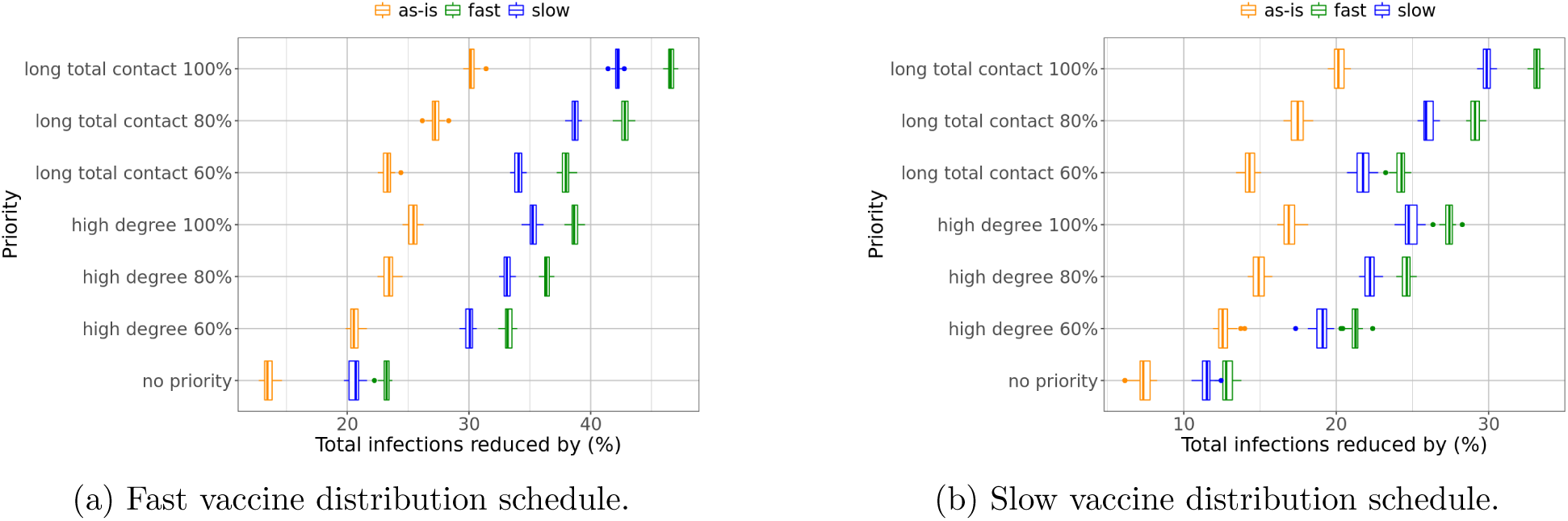
Comparison of degree and weighted degree-based strategies under (a) the fast vaccine distribution schedule; (b) the slow vaccine distribution schedule. Both can reduce infections much more than the baseline strategy. The weighted degree-based strategy outperforms the degree-based one at any prioritization level.

### 3.2 The high degree prioritization schemes are effective even when we cannot accurately estimate the degree of a node

Our results show that prioritization schemes based on degree and weighted degree (total contact time) work even when they are not accurately estimated. Specifically, even when we can only estimate the degree for 60% of the nodes (as being in the first quartile or not), we notice significant improvement in the overall control of the pandemic. This is highlighted in Figure 8, where we compare degree-based schemes of various accuracies with the age-based scheme and show improvement even at lower levels of accuracy. For example, consider infection reduction in Figure 8a: targeting high degree people with only 60% accuracy improves the reduction from 10% by the age-based strategy to 20% (with no relaxation), from 15% to 30% (with slow relaxation), or from 17.5% to 33% (with fast relaxation). Consider mortality reduction in Figure 8b: degree-based strategy with 60% accuracy improves the reduction from 17.5% to 20% (with no relaxation), from 23% to 27.5% (with slow relaxation), or from 26% to 32% (with fast relaxation). In fact, these strategies require neither knowledge of the exact degree of each person, nor that of the complete ranking of people by degree. They only depend on knowing which nodes have high degrees (are in the top quartile); they are tolerant to a certain amount of inaccuracy.

**Figure 8:**
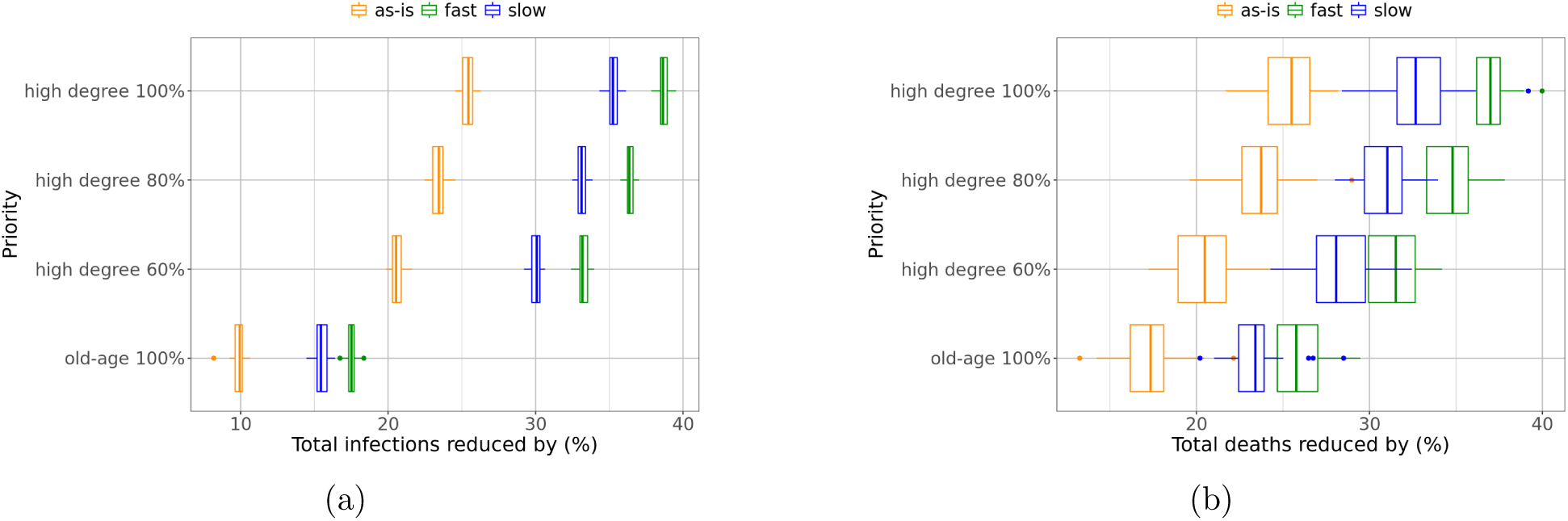
Even with lower (80% or 60%) accuracy in identifying and vaccinating high degree people, this strategy is still much more effective than the age-based strategy in (a) reducing infections, as well as (b) reducing mortality.

### 3.3 Effectiveness when social distancing measures are relaxed

The effectiveness of degree-based strategies holds in three hypothetical scenarios for social distancing: one in which there is no relaxation, and the other two wherein social distancing is progressively relaxed 5 or 7 months from now. Our results show that the value of these prioritization schemes is even higher when social distancing measures are relaxed quickly. Recall in Figure 2 we observe that, with no relaxation, the degree-based strategy results in another reduction of 85K infections and an additional reduction of 900 mortality, compared to the age-based strategy. In Figure 9, we find that with relaxation of NPIs, the degree-based strategy can reduce even more infections (152K with slow relaxation and 192K with fast relaxation) and more mortality (1.3K with slow relaxation and 1.5K with fast relaxation). These observations highlight the importance of vaccination prioritization if the current NPIs are relaxed, which will likely happen as vaccines get distributed.

**Figure 9:**
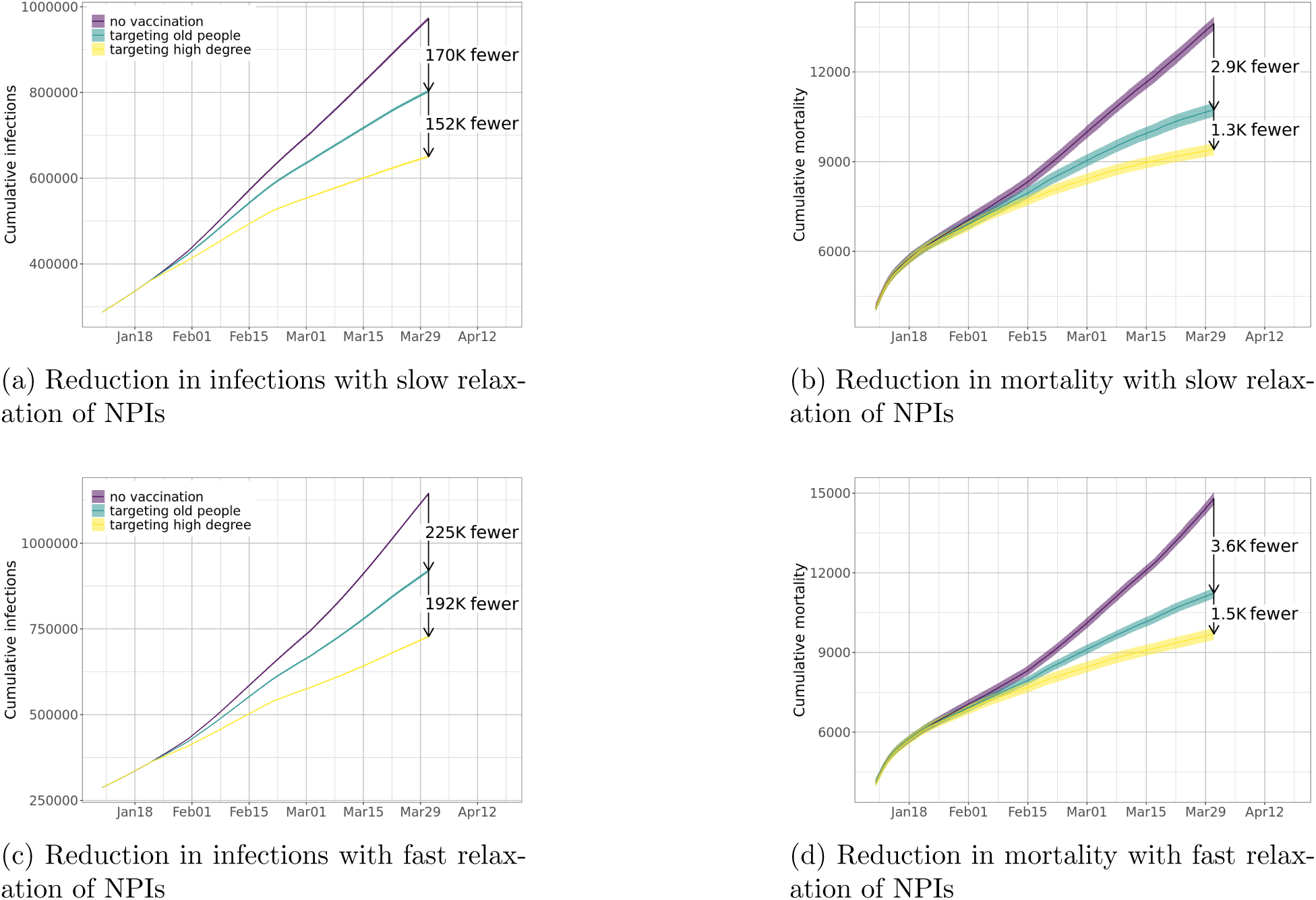
Reductions in infections and mortality from degree-based allocation strategies are even larger when NPIs are relaxed when compared to the age-based schemes. This figure shows reductions in (a) infections and (b) mortality with slow relaxation of NPIs; and reductions in (c) infections and (d) mortality with fast relaxation of NPIs.

### 3.4 Effectiveness with low efficacy vaccines

We have assumed that vaccines have 90% efficacy regarding protection against infection (*e_I_*). Our results also hold when the vaccine efficacy is lower than that of the current Pfizer and Moderna vaccines. We study this for two reasons: (*i*) there is an ongoing discussion about giving just one dose of these vaccines which may result in lower efficacy (about 50%) or approving a low efficacy vaccine^2^, and (*ii*) most other vaccines under development are traditional vaccines and may also have a lower efficacy.

To this end we study the degree-based strategies assuming 50% vaccine efficacy. In Figure 10, we show that while the reduction in infections decreases with low efficacy vaccines, it is still significant. For example, under no NPI relaxation and with the fast vaccine distribution schedule, a degree-based strategy with 60% accuracy can reduce infections by about 12.5% with *e_I_* = 50%, compared to 20% with *e_I_* = 90%. Under slow relaxation and with the slow vaccine distribution schedule, the reduction in infections is about 10% with *e_I_* = 50%, compared to 18% with *e_I_* = 90%. In Figure 11, we use epidemic curves to show reductions from the no vaccination scenario by vaccinating high degree people with 80% accuracy (*high degree 80%* in Figure 10) with both high vaccine efficacy and low efficacy, assuming slow relaxation of NPIs. We find that even with 50% efficacy, the degree-based strategy can reduce infections by 202K, hospitalizations by 13.7K, and mortality by 3.4K, by the end of March 2021, just in Virginia.

**Figure 10:**
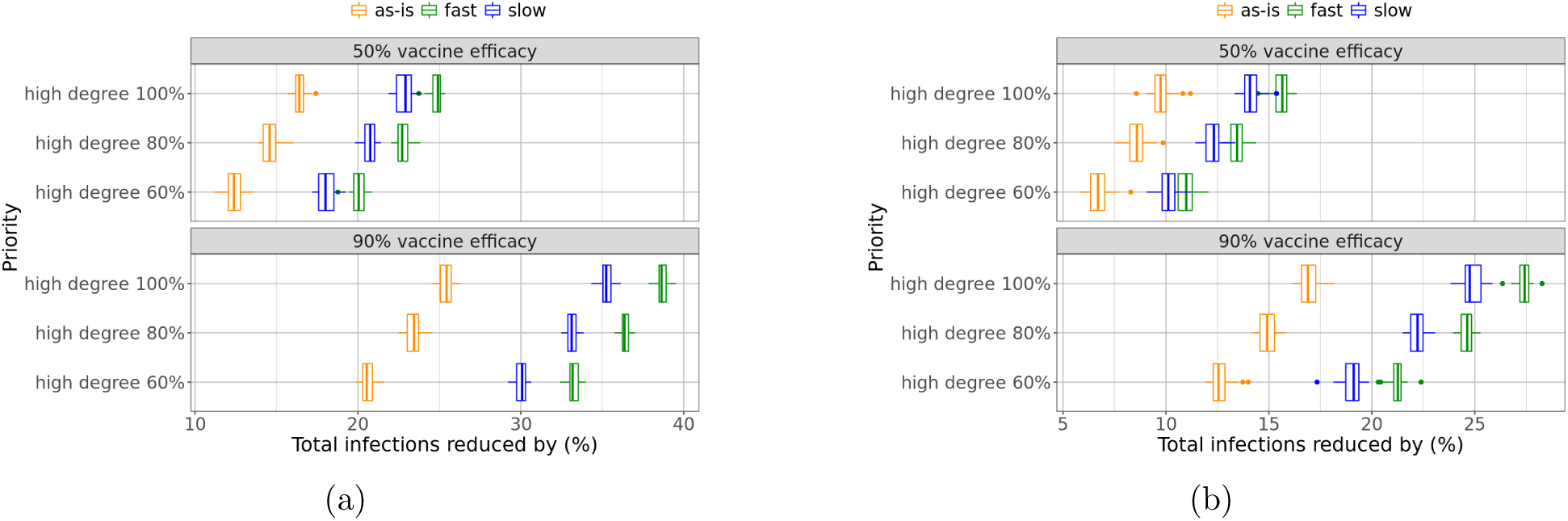
Comparison of effectiveness of degree-based strategies when vaccine efficacy is high (90%) and when it is low (50%), assuming (a) a fast distribution of vaccine schedule, or (b) a slow distribution of vaccine schedule. In both cases the effectiveness of vaccination becomes smaller with a lower vaccine efficacy, but the degree-based vaccination can still reduce infections significantly.

**Figure 11:**
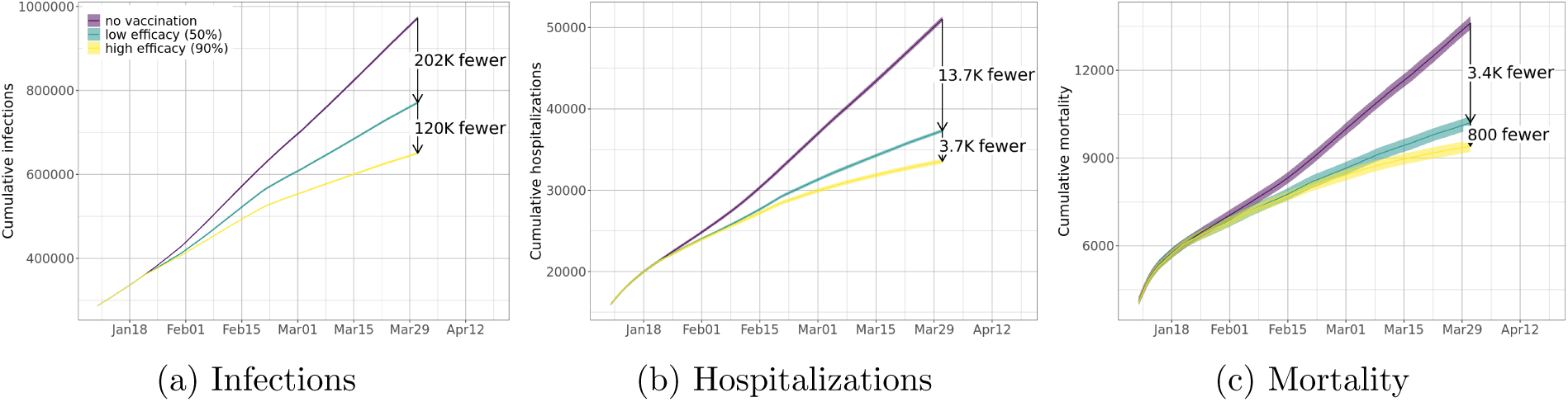
Even with a lower vaccine efficacy, a degree-based scheme can already significantly reduce (a) total infections, (b) total hospitalizations, and (c) total mortality. Numbers in the plots show total reductions up to the end of March 2021.

## 4 Discussion

These results are obtained using a realistic, data-driven and highly resolved agent-based model and individual-based social contact network of the Commonwealth of Virginia. The agent-based model represents individual-level activities that are spatially explicit. The model represents the Commonwealth-built infrastructure in great detail and uses this to develop a realistic social contact network. This allows us to: (*i*) capture details of within-host disease progression, as well as between-host transmission, including the impact of vaccines, (*ii*) model the complicated set of interventions that are currently in play, (*iii*) represent network-based vaccine prioritization schemes, (*iv*) represent the expected vaccine deployment schedule, including the expected mix of vaccine efficacy against infection, severe illness, and onward transmission estimates, (*v*) incorporate current surveillance data, and (*vi*) study counter-factual and hypothetical scenarios, such as a steady relaxation of social distancing measures. *This is the first study we know of that accounts for all of these components, not just for COVID-19, but for any infectious disease outbreak*.

The efficacy of the proposed policy is based on two important assumptions: (*i*) the synthetic contact network is a realistic representative of the real-world social contact world, and (*ii*) NPI-induced contact ‘thinning’ is applied homogeneously across the population. While the structural metrics may vary over time, we show the results are fairly robust to mis-identification of high degree individuals. We believe both these assumptions hold and discuss this in more detail below. Further discussion on this topic can be found in the Appendix, where we describe how our networks are synthesized, their structural properties, and the way the pandemic is simulated.

The potential efficacy of degree-based heuristics has been discussed in several earlier papers— this includes both provable analyses on different random graph models (under mean field assumptions in some cases), e.g., [3, 10, 51], and empirical analysis in various real world networks, e.g., [3, 22, 73]; a notion of weighted degree is also considered in [22]. However, it is important to note that these results are *not directly applicable* in our context for the following reasons: (*i*) many of the theoretical results show the efficacy of these methods for simple power law-type models – the networks we generate are similar to power law networks, but with a very different exponent; additionally the network exhibits other features of social networks (local clustering, low diameter, and relatively high expansion) and (*ii*) many of the results are shown when vaccines are applied at the start of the epidemic process, and the results do not say anything of what happens when the vaccine is applied temporally – this is important, because the temporal epidemic process infects individuals, thereby changing the network structure substantially, including the application of NPIs.

Nevertheless, the intuition behind the efficacy of such methods is simply stated as follows: vaccinating high degree nodes not only protects them, but also confers a higher level of indirect protection on their neighbors as they interact with many individuals who might themselves be conferred similar protection. Our data-driven approach shows, in fact, that real-world social net-works have sufficient nodes of high degree to ensure that such heuristics are effective. Note that, by virtue of degree bias in social networks, even traditional approaches such as contact tracing will lead us to high degree individuals. The proposed approach makes identifying these individuals as a proactive, rather than reactive, step in infection control. It is important to note, however, that just the presence of high degree nodes does not guarantee that degree-based heuristics would work. See Section E for further discussion.

As discussed earlier, identification of nodes with high degrees can be done in multiple ways, including using digital apps that have been deployed for contact tracing, interviewing individuals, and identifying typical job categories or other demographic attributes that entail higher social interactions. Further, even when other prioritization schemes are considered, one can use high social contact to further prioritize the distribution. For example, when distributing vaccines based on age, one can further subselect individuals with higher social contact in the case of limited supply. Our results suggest that degree-based prioritization should be considered by larger and resource-poor countries to quickly bend the epidemic curve and reopen the economy. The benefits of the proposed degree-based prioritization are so significant that even a partially successful campaign will likely have a large impact.

## 5 Conclusions and Limitations

We present an analysis of various vaccine prioritization strategies based on demographic attributes, occupation, and structural attributes of social contact networks. Our results show that vaccine prioritization schemes based on network degrees and total contact time can provide significant reductions in incidence, mortality, and hospitalizations. The results hold even for low efficacy vaccines and even when degrees and contact networks are estimated only approximately. Network-based prioritization is often more than twice as effective as other strategies. The results suggest that such methods should be considered when vaccines are available in limited supply; the benefits are likely to be greater in resource-poor and highly populated regions of the world. While individualized policies aimed at minimizing mortality do exist (e.g., comorbidities) and are part of the phased approach, lack of technology thus far had made it difficult to ‘individualize’ policies targeted on minimizing transmission. The advantage of our approach is in leveraging the mechanistic and network-based understanding of disease spread, and creating priority categories that cut across age, risk, and other demographic characteristics.

The study has a number of limitations, stated below. First, our network has been developed with a large number of data sources, and a number of assumptions have been made in constructing the networks, including travel patterns, distance traveled, etc. These modeling assumptions might affect the efficacy of the network-based strategies. To mitigate this, we have carried out extensive validation and assessed the impact of the uncertainty in some of the modeling parameters on the network structure. Our results indicate that the network structure is fairly robust. Second, the nodes initially infected were based on spatio-temporal and age distributions in publicly available data, but not on any network properties. If the majority of high degree nodes have already been infected and recovered, the effectiveness of targeting them for vaccination would be reduced. However, since the current vaccination policies are not based on serostatus, preferentially vaccinating higher degree individuals will still be beneficial. Furthermore, even afetr accounting for testing rates, a number of resource poor countries with large populations have so far had a relatively small outbreak. This means without vaccinations, these countries are likely to see a surge when normal worldwide travel and economic activity is resumed. The strategy is also potentially advantageous in the presence of novel variants, which may escape natural immunity. See [4, 55] for further discussion on this issue. In particular in [55], the authors point out that high degree nodes could have been infected early on in the pandemic but can pose challenges if they re-enter the pool due to waning immunity or lower immunity to new strains. This makes identifying and vaccinating high degree nodes important, even if they have been infected earlier.

Third, our base scenario has made assumptions regarding the background interventions in place. These are best estimates. Fourth, assuming that a vaccinated node gets infected, we assume that they can transmit like any other node (of course, they have a very small chance of being infected). Fifth, our results depend on estimating the degrees and weighted degrees of nodes. While we have shown that the results are robust to mis-estimation, the overall efficacy of the scheme does depend on the ability to infer these degrees.

Increasing compliance among some high degree individuals may be difficult; nevertheless, the results under such conditions will be more similar to one where the vaccine has lower efficacy and/or under degree mis-estimation. Further, when such high degree individuals are identified and vaccinated, they may themselves turn into influencers in their local community, much like the phenomenon observed on online social networks. This is a topic for immediately subsequent work. Ultimately, we believe that one can develop more comprehensive prioritization strategies that combine proposed metrics with serostatus, hesitancy surveys, and other static demographic variables to optimally reduce disease incidence and mortality.

## Data Availability

The data can be shared upon request

## Acknowledgments

The authors would like to thank members of the Biocomplexity COVID-19 Response Team and the Network Systems Science and Advanced Computing (NSSAC) Division for their thoughtful comments and suggestions related to epidemic modeling and response support. We thank members of the Biocomplexity Institute and Initiative, University of Virginia, Paul Eastham, Jeff Shaman, Chadi Saad-Roy, Theo Gibbs and Adam Sadilek for useful discussion and suggestions. This work was partially supported by National Institutes of Health (NIH) Grant 1R01GM109718, NSF BIG DATA Grant IIS-1633028, NSF Grant No.: OAC-1916805, NSF Expeditions in Computing Grant CCF-1918656, CCF-1917819, NSF RAPID CNS-2028004, NSF RAPID OAC-2027541, US Centers for Disease Control and Prevention 75D30119C05935, University of Virginia Strategic Investment Fund award number SIF160, Google Grant, and Defense Threat Reduction Agency (DTRA) under Contract No. HDTRA1-19-D-0007. Any opinions, findings, and conclusions or recommendations expressed in this material are those of the author(s) and do not necessarily reflect the views of the funding agencies.

## A Models and Methods

We study vaccine allocation strategies using agent-based simulations, which compute COVID-19 disease spread in a population (e.g. Virginia) through a social contact network. In this section, we describe a synthetic contact network of Virginia and the agent-based simulation model of COVID-19. Then we describe vaccine allocations based on different prioritizations, especially strategies targeting high degree people in the population. The overall framework is described in Figure 13.

**Figure 12:**
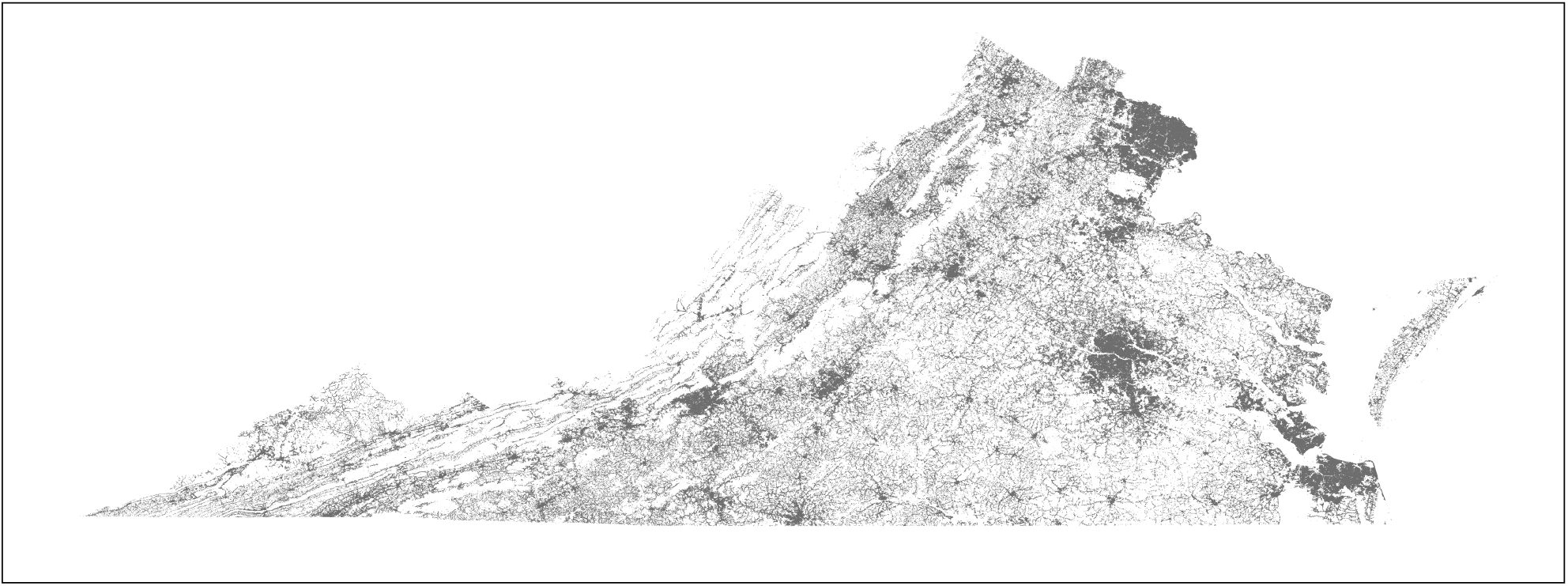
The illustration shows the locations for the synthetic population of Virginia used in this paper. Each location’s centroid (longitude and latitude) is shown as a point.

**Figure 13:**
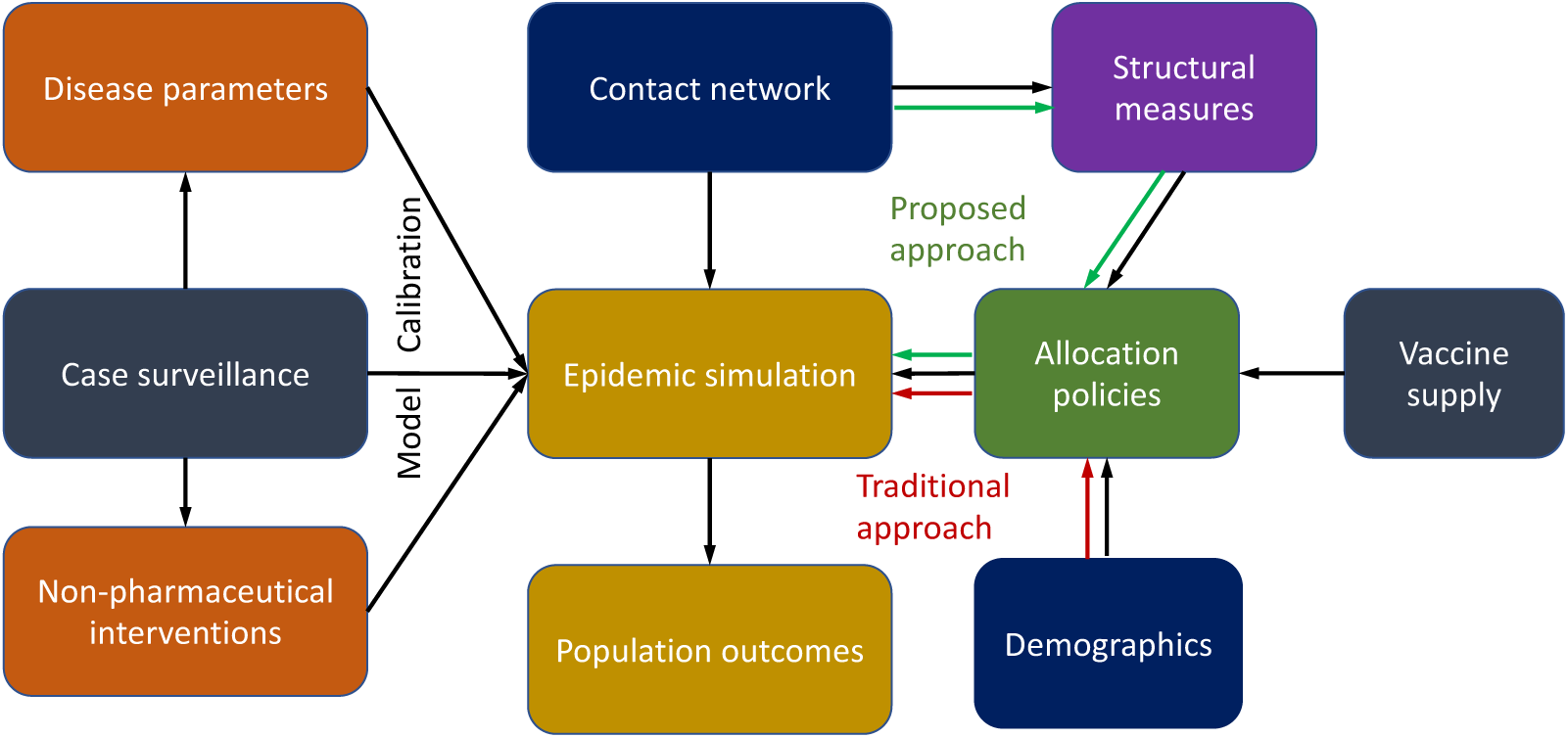
The overall data driven framework used to study various prioritization strategies The figure shows schematically how real world data is used to drive the modeling and analysis process. This provides a realistic context for the underlying the simulations. For instance, it takes into account the current disease prevalence, vaccine schedule, efficacy, NPIs into account to evaluate the strategies.

### A.1 Generating synthetic populations and networks

A synthetic population of a region may be regarded as a digital twin of the real population of that region. Here we provide a compact summary of the model and the methodology behind constructing synthetic populations and their contact networks in the case of the US; see [47] for details. Our work builds on earlier techniques for a first principles approach for constructing synthetic populations [7, 23, 24]. These populations and networks are central to the EpiHiper simulation model.

To construct a population for a *geographic region R* (e.g., Virginia), we first choose a collection of *person attributes* from a set 𝒟 (e.g., age, gender, and employment status) and a set 𝒯*_A_* of *activity types* (e.g., `Home`, `Work`, `Shopping`, `Other`, and `School`). The precise choices of 𝒟 and 𝒯*_A_* are guided by the particular scenarios or analyses the population will serve. Described at a high level, we (*i*) construct people and places, (*ii*) assign activity sequences to people, (*iii*) map each activity for each person to a location (including the time of the visit), and (*iv*) from this, we derive a contact network using co-occupancy to infer edges. The construction is broken down in a sequence of steps outlined as follows.

Using *iterative proportional fitting* (IPF) [8, 20] the **base population** model constructs a set of individuals 𝒫 where each person has assigned demographic attributes from 𝒟. By design, this ensures that 𝒫 matches the actual distributions and Public Use Microdata Sample (PUMS) data from the US Census [64], which is the input data for the model. Additionally, this model partitions 𝒫 into a set ℋ of *households*, where the notion of household encompasses the traditional notion of “family”, but also any other subset of individuals residing in the same *dwelling unit* (e.g., dormitories, army barracks, or prisons).

After household assignment, each individual *p* ∈ 𝒫 is assigned a week-long activity sequence *α*(𝒫) = (*a_i,p_*)*_i_* where each *activity a_i,p_* has a *start time*, a *duration*, and an *activity type* from 𝒜. Data sources used for this step include National Household Travel Survey (NHTS) [66], American Time Use Survey (ATUS) [65] and Multinational Time Use Study (MTUS) [61]; these sources are fused to form consistent, week-long activity sequences. We write 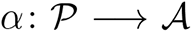 for the mapping assigned to each person. For this construction, we use Fitted Values Matching (FVM) for adults [39], and Classification And Regression Tree (CART) for children (see, e.g., [11]).

The **location model** constructs a set of spatially embedded locations ℓ consisting of *residence locations* where households live, and activity locations where people conduct their non-`Home` activities. This construction is highly granular and is rooted in data such as the MS Building data [44], HERE/NAVTEQ data [33] for points-of-interest (POIs) and land-use classifications, National Center for Education Statistics (NCES) [49] data for public schools, as well as LandScan^3^, OpenStreetMap^4^, and Gridded Populations of the World (GPW) v4^5^. A point plot of the locations of Virginia is shown in Figure 12.

For each person *p ∈ P*, the **location assignment model** assigns a location ℓ = ℓ(*a_i_*) to each of their activities *a_i_*. We denote the sequence of locations visited by 𝒫 as *λ_p_* = (*λ_i_*)*_p_*. The location assignment model uses American Community Survey (ACS) commute flow data [63] to assign the target county *c* for `Work` activities, and a particular location randomly within *c* work weights assigned to each location in *c*. School activity locations are assigned based on NCES data, with remaining activities anchored near home and work locations.

Finally, the **contact network model** uses the location assignment to derive the bipartite *people location graph G_P L_* with vertex sets *V*_1_ = 𝒫 and *V*_2_ = ℓ and a labeled edge (*p, ℓ*) whenever 𝒫 visits ℓ where the label includes activity type, time for start of visit, and duration of visit. From this, we derive the list of visitors to each location and the *co-location graph G*_max_ with vertex set 𝒫 and edges all *e* = (*p, p^’^*) for people *p* and *p^’^* that are simultaneously present at the same location. Merely being present at a location at the same time does not imply a contact, and sub-location contact modeling is applied at each location to determine which of the edges of *G*_max_ should be retained to form the *contact network G* which is also referred to as the *person-person contact network* and denoted as by *G_P P_* (rather than simply *G*) to make this explicit. In this work, we use a random graph model referred to as the *Min/Max/alpha model* at each location to obtain *G*. Let ℓ be a location and let *N* = *N_l_* denote the maximal number of simultaneous visits to ℓ. Define the function 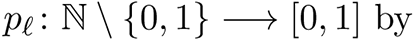

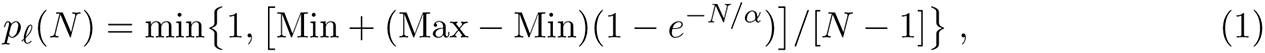

where Min *<* Max are non-negative numbers and *α >* 0. Given *p* = *p_ℓ_*(*N*) one samples from this random graph model in the same manner as for the standard model *G_n,p_* by independently at random applying to each edge *e* the probability *p* corresponding to the location ℓ where *e* ∈ *G*_max_ originate. Thus the parameters Min and Max bound the degree of each vertex locally at ℓ (in expectation) for each visit; note, however, that the degree of person *p* in the resulting graph *G* is the accumulation of degrees across their trajectory to locations visited while executing their activity sequence. Thus the choices of Min, Max and *α* will induce the degree of each vertex in a bottom-up manner, see [47] for full details. Finally, for the applications and scenarios of this paper, we project from *G*, the week-long contact network, to *G*_Wednesday_, representing the contact network on a “typical day”.

We will use agent-based network models to study the epidemic process in this paper. Agent-based networked models (sometimes just called agent-based models) extend metapopulation models by explicitly capturing the interaction structure of the underlying populations. In this class of models, epidemic dynamics are modeled as a diffusive process on a specific undirected person-person contact network *G*(*V, E*) = *G_P P_* (*V, E*) on a population *V* (see Section A.1 for precise definitions) – the existence of an edge *e* = (*u, v*) ∈ *E* implies that individuals (also referred to as nodes) *u, v ∈ V* come into contact^6^. Let *N* (*v*) denote the set of neighbors of *v*.

#### Epidemic process over networks

The SIR model on the person-person contact network *G* is a dynamical process in which each node is in one of three states: *S*, *I* or *R*. Infection can potentially spread from *u* to *v* along edge *e* = (*u, v*) with a probability of *β*(*e, t*) at time *t* after *u* becomes infected, conditional on node *v* remaining uninfected until time *t*— this is a discrete version of the rate of infection for the classical compartmental mass action models discussed earlier. We let *I*(*t*) denote the set of nodes that become infected at time *t*. The (random) subset of edges on which the infections spread is referred to as a *dendrogram*. This dynamical system starts with a configuration in which there are one or more nodes in state I and reaches a fixed point in which all nodes are in states S or R. *In our simulations, the disease models are significantly more complicated than simple SIR processes; this is described in the* Appendix in *Section G*.

### A.2 Interventions and vaccine allocation policies

Interventions are implemented to inhibit disease transmission. Interventions can be thought of as individual behavioral adaptations or policy mandated changes, such as closing certain facilities or reducing their capacity. Of course, policies also lead to further behavioral adaptations. Our agent-based models have a rich set of interventions implemented. The specific ones we use in the study are detailed in Section F. In our simulations, all policy changes and behavioral adaptations except vaccine uptake can be seen as processes that continually and adaptively change the social contact network and disease transmission parameters.

Here we focus on policy concerning vaccine allocation. Given a schedule *S* that specifies the amount of each vaccine available at each time, and the characteristics of the vaccines, a prioritization scheme is a policy that assigns at each time period the individuals (nodes) that are to be vaccinated. In other words, a vaccine prioritization scheme can be thought of as a Markov Decision process – at each time step we know the current state of the system and the available vaccines, and we need to decide who gets the vaccine. In this paper, we only consider non-adaptive policies – i.e. policies that do not change how vaccines are allocated at each time step. The amount depends, of course, on the schedule. We focus on four types of policies, each of which partitions the population into priority groups using different characteristics:

1. age group,
2. occupation,
3. number of social contacts (degree), and
4. total duration of social contacts (weighted degree).
Within each type, the policies are distinguished by the fraction allocated to each priority group (further details can be found in Section 2.2).

#### The criteria for evaluation of policies

A policy’s effectiveness will be measured by comparing the total numbers of the following to a baseline case with no vaccines: (*i*) infections, (*ii*) hospitalizations, and (*iii*) deaths.

Note that the simulation output is a random variable and thus we report the empirical expectation and variance of this random variable, where the expectation is taken over all possible initializations of the stochastic process and the probabilistic transmission and intervention process. We follow the discussion in [59]. Formally, at any time *t*, the state of the system *S_t_* = (*I_t_, W_t_*), where *I_t_* and *W_t_* denote the set of infected and vaccinated nodes at time *t*. The stochastic process is started in some initial condition *S*_0_ = (*I*_0_*, W*_0_). The decision to vaccinate is done over time horizon *T* and the vaccine schedule is given by *S*. Given this for a policy *π*, its expected utility with respect to infection (criterion *i* above) is given by

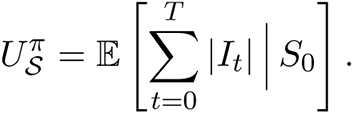

An agent-based simulation model is described in Appendix F. The COVID-19 disease model is described in Appendix G.

## B Structural Measures for Social Contact Networks

In this section, we define some popular network measures used to characterize real-world networks and use them to study the structure of the contact networks used in this work. These include measures of local connectivity such as degree and clustering coefficient, global connectivity such as diameter, *k*-core, and graph spectrum. These measures have been used in both structural and dynamical characterization of real-world networks. In particular, these measures are considered important with respect to epidemic simulations, and are frequently reported in the literature [14, 28]. Our objective is two-fold: (i) compare our networks with existing real-world networks, and (ii) use these measures to analyze the efficacy of the different control strategies applied in our work. The latter is discussed in Section E.

The construction of the synthetic contact network of Virginia is described in Appendix A.1 along with definitions of the people-location network *G_P L_* and the person-person contact network *G* = *G_P P_*. Note that this network was modeled and constructed with epidemics and disease transmission as a target. Generally, what constitutes an interaction (and thus edge) factors through physical proximity, the nature of the interaction, the nature of the dynamics studied (e.g. disease transmission) and other factors such as, for example, air circulation within a building and infection through contaminated inanimate objects. Note first that the person-person contact network *G* has 7.6*×*10^6^ nodes and 2.0*×*10^8^ edges. Moreover, *G* has a largest component of (relative) size 0.983.

### Diameter

The diameter is the length of the longest shortest path between any two vertices of the network. The diameter of *G* is diam(*G*) = 12.

### Degree distribution

The degree of a person *u* in the people-people contact network *G* is the number of different persons that *u* has contact with during a day. The average person degree in *G* is 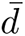 = 43.5. In the people-location network *G_P L_*, the degree of a location ℓ is the number of distinct visitors to ℓ during a day, and the degree of a person *u* is the number of distinct locations *u* visits during a day. Figure 14 shows the degree distributions in *G* and *G_P L_*. The weighted degree of a node *u* is the total time of contact with its neighbors per day. The weighted degree distribution of the network *G* is shown in Figure 15a.

**Figure 14:**
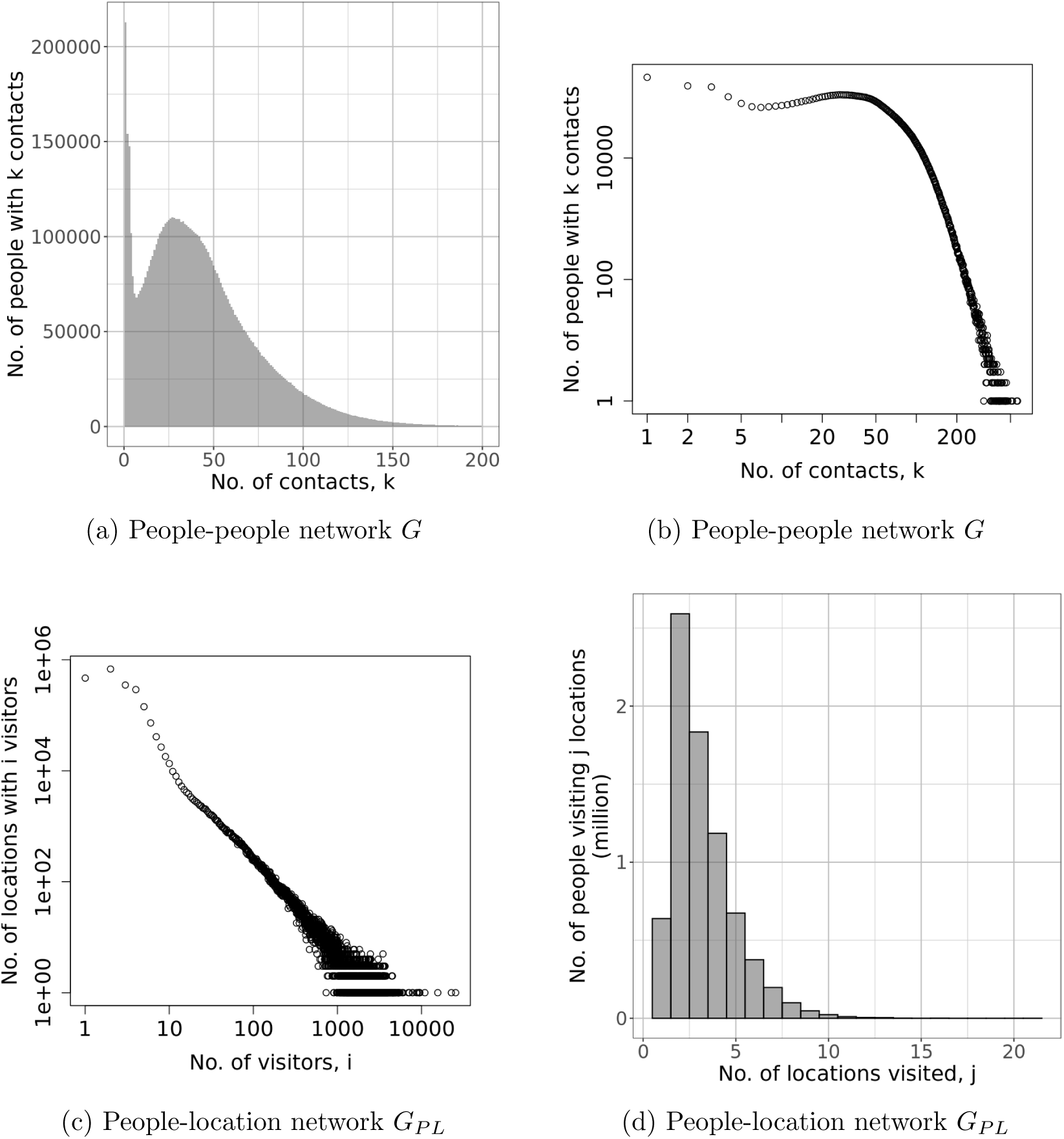
Degree distributions in people-people network *G*: (a) as a histogram in normal scale, and (b) in log-log scale showing a power-law tail with an exponent of about −7; and degree distribution in people-location network *G_PL_*: (c) of locations in log-log scale showing a power-law tail with an exponent of about −1.7, and (d) of people as a histogram in normal scale.

**Figure 15:**
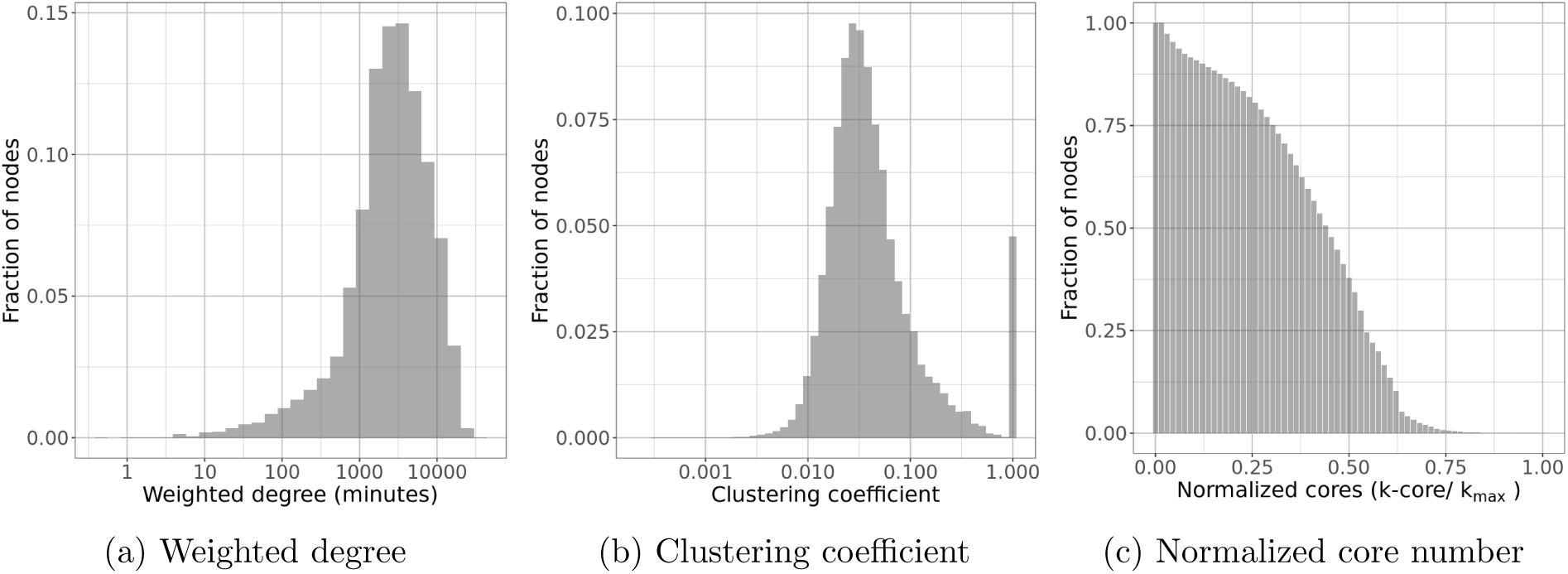
Distributions for connectivity properties of *G* here including (a) the weighted degree distribution, (b) the clustering coefficient distribution, and (c) the normalized core number distribution for *G*. of the people-people network.

### Clustering coefficient

This is a measure of the degree to which nodes in a graph tend to locally cluster together. For a node *v*, its *local clustering coefficient* is the fraction of pairs of its neighbors which have a link between them. It quantifies how close the immediate neighbors are to being a complete graph. If the local clustering coefficient is 1, it means that the node and its neighbors induce a complete graph, and if it is 0, then they induce a star graph with *v* at the center. The *average clustering coefficient* is the average of all the local clustering coefficients [70]. The average clustering coefficient of *G* is 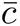 = 0.092; the clustering coefficient distribution is shown in Figure 15b. *k***-core.** A *k*-core of a graph *G* is a maximal connected subgraph of *G* in which all vertices have degree of at least *k*. Equivalently, it is one of the connected components of the subgraph of *G* formed by repeatedly deleting all vertices of degree less than *k* [71]. The *core number* of a graph is the maximum core it belongs to. The *maximum core* (max. core) of the graph is the set of all nodes with the maximum core number. It is another way of characterizing centrality of nodes. It is considered that for certain epidemiological models, the *k*-core number is a good predictor of the final outbreak size [50]. Also, the *k*-core decomposition is in many cases a good predictor of spreading efficiency. The *k*-core decomposition of *G* is shown in Figure 15c. The maximum *k* for which there is a *k*-core is 57. While there are only 1,700 nodes in the induced subgraph for the 57-core, there are approximately 35,000 nodes (around 0.5% of the total population) in the 45-core, and more than a quarter of the population belongs to the 30-core showing high global connectivity

### Graph spectrum or eigenvalues

The spectrum of a graph is the set of eigenvalues of its adjacency matrix. Also popular is the Laplacian spectrum, which is the set of eigenvalues of the Laplacian matrix of the graph. There are several works that relate spectrum, particularly the first eigenvalue of the adjacency matrix, to disease spread in SEIR-like models [27, 52]. The common result that highlights the impact of the network structure on the dynamics is that epidemics die out “quickly” if *λ*_1_(*G*) *≤ T*, where *λ*_1_(*G*) is the *spectral radius* (or the largest absolute value of an eigenvalue) of graph *G*, and *T* is a threshold that depends on the disease model. This relationship has motivated a number of works on epidemic control where the objective is to find an optimal set of nodes (or edges) to remove from the network that leads to maximum reduction in its spectral radius [57, 67, 74].

### Activity-based structural analysis

We analyzed the constructed social contact network using the structural measures described above. We considered activity induced sub-networks, where a contact edge is retained only if both individuals corresponding to that edge have the target activity assigned to them. For example, in the case of `School` network, only `School–School` edges are retained. For `School` activity, we observed an average degree of 29.7, which is between the average degrees of 13.5 and 47.3 reported for the school networks of the SocioPatterns collaboration networks [14, 28]. Our average degree for `Work` activity is 16.3. In comparison to the SocioPatterns data, this seems to be on the higher side for a typical office environment (*<* 7) but comparable to their hospital network (14.0). This is expected, since our `Work` activity includes a wide variety of workspaces that include office environment, factories, restaurants, etc. The max. core for the `School` activity network is 32. In SocioPatterns, the two schools have max. cores of 24 and 47 respectively. For the `Work` network, it is 37, while in SocioPatterns it is 11, 25 and 23 respectively for the three office networks. Again, in our case the max. core is the maximum among all locations.

## C Additional Results and Analysis

We present additional results as discussed in the paper.

### C.1 Epidemic curves with various strategies

Figures 16 to 18 show the cumulative numbers of infections, hospitalizations, and mortality in the first three months of 2021 under different vaccination schemes, assuming no relaxation of NPIs (Figure 16), slow relaxation (Figure 17), and fast relaxation (Figure 18). Note that for the age-based, essential worker-based, and degree-based schemes, we only show the moderate level of priority enforcement 80%. All curves show the uncertainty of one standard deviation above and below the mean.

**Figure 16:**
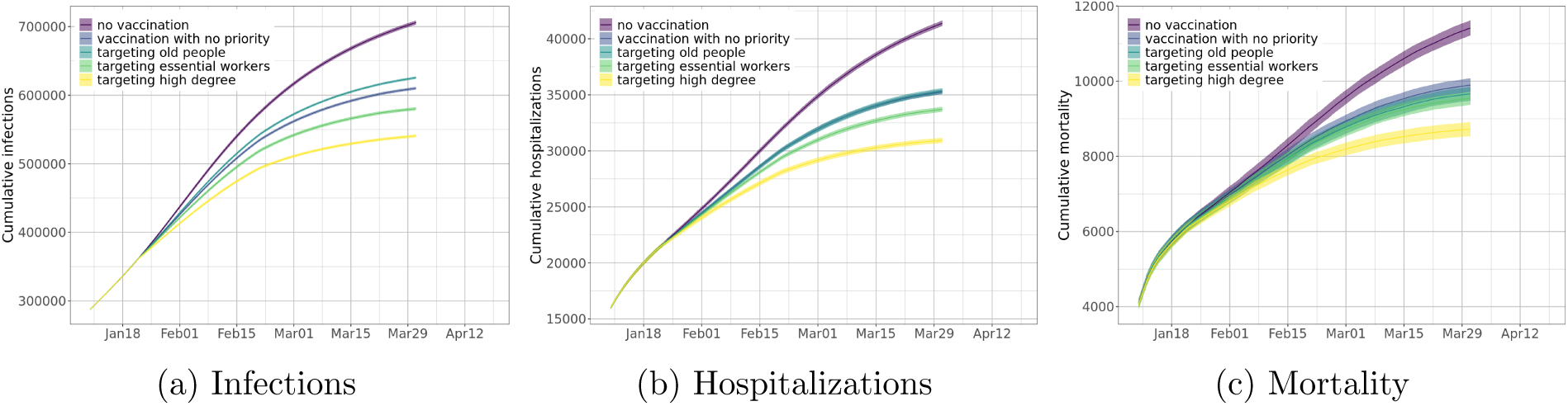
Cumulative counts under different vaccination strategies for (a) total infections, (b) total hospitalizations, and (c) total mortality, assuming current non-pharmaceutical interventions remain at the same level (*as-is*).

**Figure 17:**
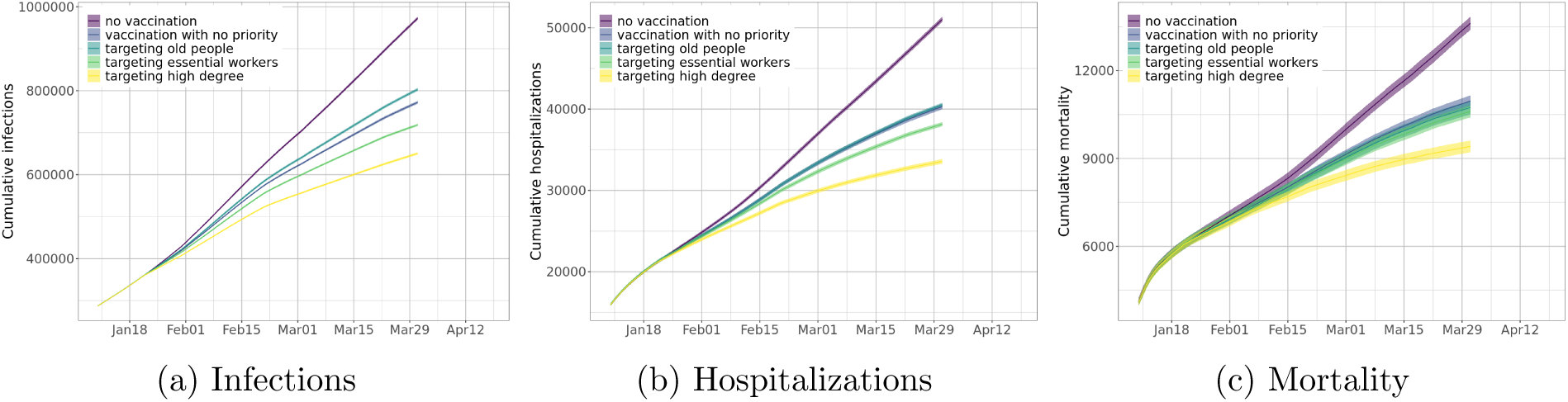
Cumulative counts under different vaccination strategies for (a) total infections, (b) total hospitalizations, and (c) total mortality, assuming slow relaxation of non-pharmaceutical interventions.

**Figure 18:**
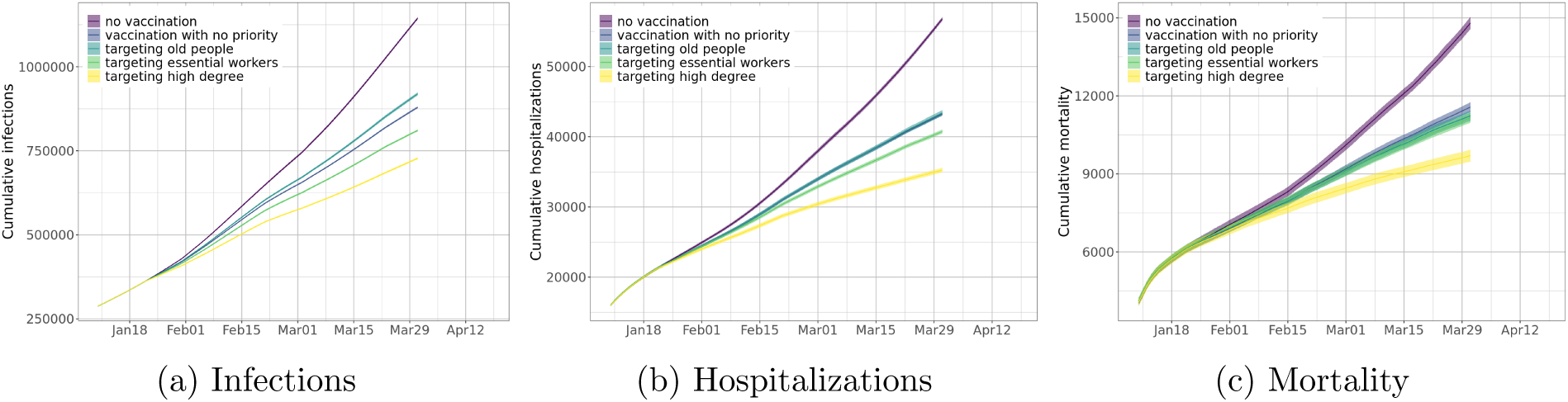
Cumulative counts under different vaccination strategies for (a) total infections, (b) total hospitalizations, and (c) total mortality, assuming fast relaxation of non-pharmaceutical interventions.

#### Infections

We find that in all three scenarios of relaxation, while vaccinations can significantly reduce infections, targeting high degree people clearly outperforms all other schemes; and the no-priority scheme performs better than the age-based scheme but not as well as the essential worker targeting scheme.

#### Hospitalizations

While the no-priority scheme outperforms the age-based scheme on reducing infections, they seem to have similar reductions on hospitalizations. This is because older people have fewer contacts on average, so vaccinating them does not decrease the connectivity of the network as much as the other schemes. But since vaccines can protect the vaccinated against severe illness (we assume *e_D_* = 50%), and the older people have a higher hospitalization ratio, so targeting them is more effective on reducing hospitalizations than on reducing infections.

#### Mortality

Due to the same protection to the vaccinated against severe illness, targeting old people seems more effective on reducing mortality than on reducing infections. Targeting essential workers is marginally better than the age-based and the no-priority schemes. The degree-based scheme, however, continue to be the most effective on reducing mortality.

We also observe that when NPIs are relaxed, vaccinations become more important. This is discussed in Section 3.3.

### C.2 Sensitivity analysis

This section shows some of the variability in network measures for a statistical design over the network construction parameters Min, Max and *α*. Specifically, we consider the hyperplane given by Min = 5, *α* = 1000 and Max *∈ {*35, 40, 45*}* with Max = 40 representing the base case used in all the simulations of this work. Accordingly, we have three graphs G-35, G-40, and G-45. As can be seen in Table 2, there is a slight increase in properties such as 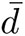, 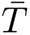, and d_max_ while the diameter drops by 1 for Max = 45. The slightly bigger change occurs in eigenvalues, but we note that the spectral gap (i.e., *λ*_1_ *− λ*_2_) remains nearly constant.

**Table 2:** Here *|V |* denotes the number of nodes, *|E|* the number of edges, 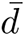the average degree, 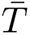 the average total contact time (unit: hour), d_max_ the maximal degree, *T*_max_ the maximal total contact time, 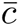 the average clustering coefficient, *k*_max_ the maximal core, *ρ*_max_ the relative size of the maximal component, diam the network diameter, and *λ*_1_ and *λ*_2_ the two largest eigenvalues.

| Network | $\vert V\vert $ | $\vert E\vert $ | $\bar{d}$ | $\bar{T}$ | $d_{\max}$ | $T_{\max}$ | $\bar{c}$ | $k_{\max}$ | $\rho_{\max}$ | diam | $\lambda_1, \lambda_2$ |
| --- | --- | --- | --- | --- | --- | --- | --- | --- | --- | --- | --- |
| G-35 | 7.6e06 | 2e+08 | 41.0 | 54.4 | 455 | 588.5 | 0.091 | 51 | 0.983 | 12 | 100.6, 87.8 |
| G-40 | 7.6e06 | 2e+08 | 43.5 | 59.2 | 565 | 1526.9 | 0.092 | 57 | 0.983 | 12 | 118.5, 100.6 |
| G-45 | 7.6e06 | 2e+08 | 49.2 | 64.4 | 636 | 655.6 | 0.096 | 66 | 0.983 | 11 | 127.1, 110.0 |

## D Analysis of the time-varying network

Each time the chosen control strategy is applied to the network, some subset of nodes are removed leading to structural changes in the network. We analyzed this evolution of the network using the various measures introduced in Section B. The network measures for each residual network are listed in Table 3. For ease of comparison, the evolution of the max. core and the spectral radius are plotted in Figure 19. Also, in Figure 20, we have plotted the distribution of core number.

**Figure 19:**
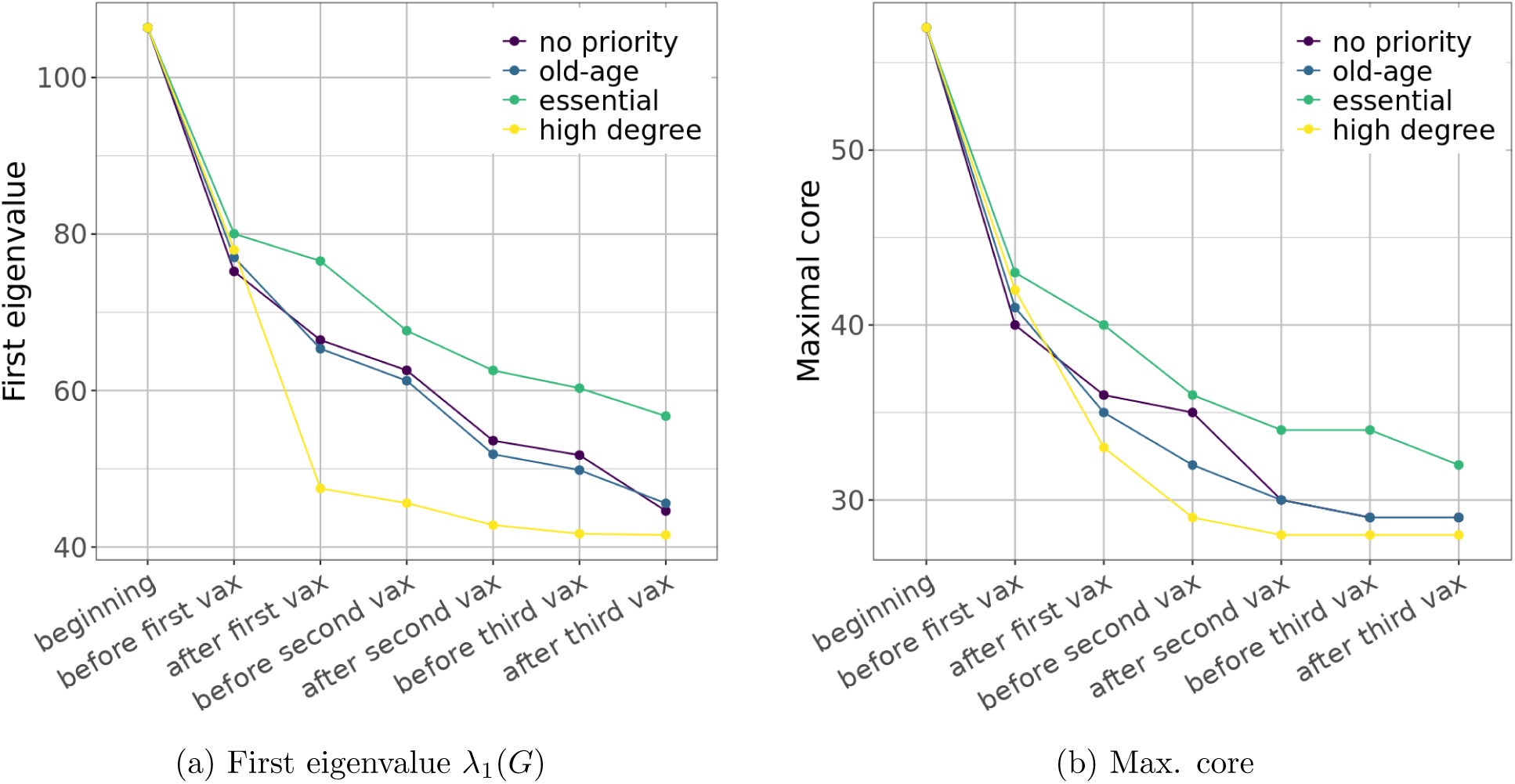
Reduction in the first eigenvalue and max. core of the network at different vaccination phases. As nodes are infected or vaccinated, we remove edges incident on them from the contact network, to form sparser and sparser *snapshot* networks. We consider such snapshots under different prioritization strategies, assuming no relaxation of NPIs and 100% distribution. After the first round of vaccination, the degree-based strategy reduces the first eigenvalue and the max. core the most as compared to other strategies.

**Figure 20:**
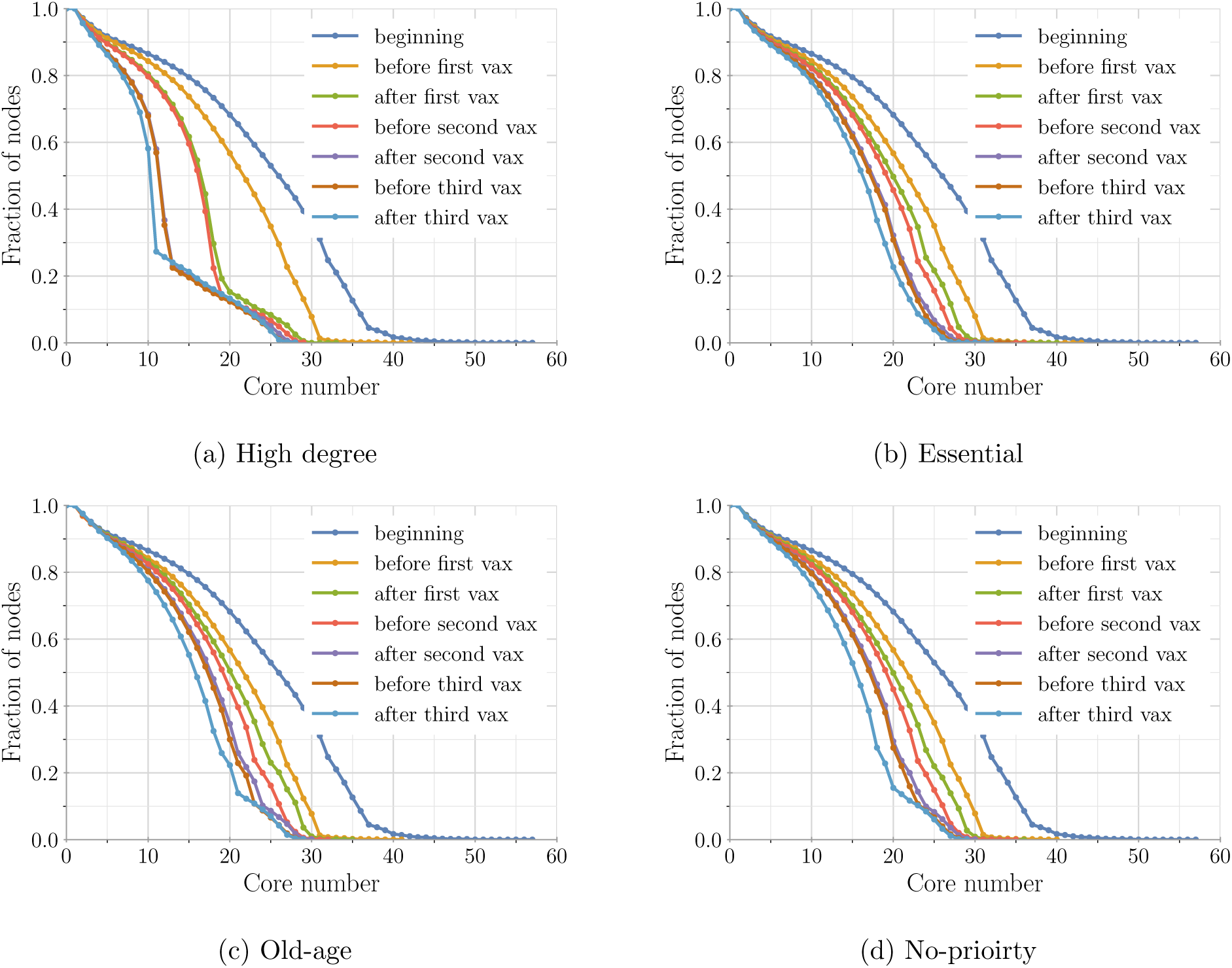
The evolution of *k*-core distribution in the snapshot networks extracted at various stages of the simulation. Different vaccination prioritization strategies are implemented in the four subfigures. Note that these snapshot networks are the same ones used in Figure 19 and Table 3.

**Table 3:** Properties of the temporal snapshots of the social contact network before and after each application of vaccination. The “Strategy” column refers to the vaccination prioritization. Each line corresponds to a snapshot network extracted from the contact network by removing edges incident on infected nodes and vaccinated nodes. These snapshot networks are the same ones used in Figure 19.

| Strategy | Snapshot | Nodes | Avg. deg. | Max. deg. | Max. contact hrs. | Avg. clust. coeff. | Max. core ($k_{\max}$) | Giant comp. | Diameter | Top eigenvalues |
| --- | --- | --- | --- | --- | --- | --- | --- | --- | --- | --- |
| none | beginning | 7605430 | 43.5 | 565 | 3053.7 | 0.092 | 57 | 0.983 | 12 | 118.5,100.6 |
| high degree | before first vax | 6306549 | 35.8 | 400 | 2161.8 | 0.086 | 42 | 0.984 | 12 | 87.8,79.5 |
| essential | before first vax | 6305803 | 35.8 | 507 | 2165.2 | 0.086 | 43 | 0.984 | 13 | 89.7,80.5 |
| old-age | before first vax | 6301976 | 35.7 | 512 | 1664.6 | 0.086 | 41 | 0.983 | 13 | 85.1,79.0 |
| no-priority | before first vax | 6305637 | 35.8 | 504 | 1948.7 | 0.086 | 40 | 0.983 | 12 | 83.2,77.0 |
| high degree | after first vax | 5719119 | 26.6 | 374 | 1673.4 | 0.09 | 33 | 0.981 | 13 | 48.9,46.2 |
| essential | after first vax | 5704209 | 32.6 | 492 | 1999.0 | 0.082 | 40 | 0.984 | 13 | 83.6,74.3 |
| old-age | after first vax | 5679458 | 33.1 | 506 | 1647.1 | 0.085 | 35 | 0.987 | 12 | 72.4,65.0 |
| no-priority | after first vax | 5698684 | 32.8 | 494 | 1847.5 | 0.082 | 36 | 0.985 | 12 | 73.8,67.5 |
| high degree | before second vax | 5605355 | 25.7 | 363 | 1278.2 | 0.09 | 29 | 0.981 | 13 | 47.4,44.9 |
| essential | before second vax | 5563558 | 31.3 | 457 | 1292.0 | 0.082 | 36 | 0.983 | 13 | 71.0,64.7 |
| old-age | before second vax | 5500145 | 31.3 | 487 | 1287.9 | 0.086 | 32 | 0.986 | 13 | 67.0,61.4 |
| no-priority | before second vax | 5534739 | 31.1 | 480 | 1350.5 | 0.083 | 35 | 0.984 | 13 | 67.5,62.6 |
| high degree | after second vax | 5059583 | 19.4 | 179 | 1259.7 | 0.097 | 28 | 0.977 | 14 | 45.3,42.4 |
| essential | after second vax | 4961207 | 28.4 | 435 | 1292.0 | 0.078 | 34 | 0.984 | 12 | 65.2,58.3 |
| old-age | after second vax | 4888575 | 28.8 | 380 | 1287.0 | 0.086 | 30 | 0.989 | 13 | 56.7,52.0 |
| no-priority | after second vax | 4929293 | 28.3 | 363 | 1347.9 | 0.079 | 30 | 0.986 | 13 | 58.2,51.3 |
| high degree | before third vax | 5022947 | 19.1 | 172 | 1259.7 | 0.098 | 28 | 0.977 | 14 | 43.9,41.3 |
| essential | before third vax | 4914302 | 28.0 | 423 | 1232.2 | 0.078 | 34 | 0.983 | 13 | 64.0,60.2 |
| old-age | before third vax | 4813271 | 28.1 | 370 | 1227.7 | 0.087 | 29 | 0.988 | 13 | 54.1,49.8 |
| no-priority | before third vax | 4864470 | 27.7 | 360 | 1331.1 | 0.079 | 29 | 0.986 | 13 | 56.4,51.6 |
| high degree | after third vax | 4520360 | 18.3 | 169 | 1259.7 | 0.092 | 28 | 0.98 | 14 | 43.7,41.1 |
| essential | after third vax | 4533299 | 26.3 | 273 | 1219.0 | 0.076 | 32 | 0.983 | 12 | 61.3,57.3 |
| old-age | after third vax | 4212668 | 25.8 | 361 | 1227.7 | 0.088 | 29 | 0.991 | 13 | 47.8,45.5 |
| no-priority | after third vax | 4256026 | 25.0 | 349 | 1331.1 | 0.074 | 29 | 0.988 | 13 | 46.8,44.3 |

**Table 4:** EpiHiper state values of nodes and edges

| Object | Value | Access | Description |
| --- | --- | --- | --- |
| system | time | r | the current time (mapped from iteration number/tick) |
| node | id | r | PID of node |
| node | infectivity | rw | infectivity scaling factor |
| node | susceptibility | rw | susceptibility scaling factor |
| node | healthState | rw | health state |
| node | nodeTrait[traitName] | rw | value of nodeTrait[traitName] of node |
| edge | sourceID | r | source vertex ID of edge |
| edge | targetID | r | target vertex ID of edge |
| edge | sourceActivity | r | activity of source of edge |
| edge | targetActivity | r | activity of target of edge |
| edge | active | rw | active flag of edge |
| edge | weight | rw | weight of edge |
| edge | edgeTrait[traitName] | rw | value of edgeTrait[traitName] of edge |
| variable | name | rw | a user-defined numerical variable referenced by name |

The measures that are generally affected by vaccination are max. degree, avg. degree, max. core and eigenvalues. Diameter and size of giant component remain little changed over time. We note that among all strategies, the most significant change in network properties is observed for the degree-based strategy. The efficacy of targetting old population and targetting essential workers are comparable to random vaccinations with respect to these measures.

## E Why do social interaction-based heuristics work and how can they be implemented

We briefly discuss the reasons why the degree- and weighted degree-based heuristics work. We also briefly discuss the role of digital devices in estimating degrees of individuals.

### Why do degree-based heuristics work

The efficacy of degree-based heuristics has been discussed in several earlier papers, e.g., [3,10,22,51,73], as discussed in Section H. This phenomenon is also well understood in certain random graph models [10, 51]. Pastor-Satorras et al. [51] derive an immunization threshold in terms of the fraction of high degree nodes that are immunized through an analysis that relies on a degree-based mean field (DBMF) assumption. This is derived more rigorously by Bollobas et al. [10], who show that there is a threshold fraction of high degree nodes whose removal shatters the graph—this can be viewed as an immunization strategy in their random graph model for the case when the transmission probability 𝒫 = 1.

However, it is important to note that the theoretical results are *not directly applicable* in our context for the following reasons: (*i*) many of the theoretical results show the efficacy of these methods for power law networks – the networks we generate are similar to power law networks but with a very different exponent; and (*ii*) many of the results are shown when vaccines are applied at the start of the epidemic process, and the results do not say anything of what happens when the vaccine is applied temporally – this is important, because the temporal epidemic process infects individuals already and thus changes the network structure substantially, including the application of the non-pharmaceutical interventions.

As nodes are infected or vaccinated, they are either not susceptible or less susceptible (i.e. lower probability of getting infected). Consider the network structural changes due to such dynamics. For simplicity, we remove all edges incident on nodes who are or have been infected and those who are vaccinated, to form a sparser *snapshot* network at different time point, especially right before and right after each mass vaccination. In Figure 21, we compare the degree distributions of such snapshot networks from age-based and degree-based vaccinations, before and after each vaccination. The networks are extracted from a simulation run under as-is scenario with fast vaccine distribution and 100% prioritization. Before the first vaccination, the snapshot network from both strategies have the same degree distribution. After the vaccination, since more edges are removed with the degree-based strategy, the snapshot network has more low degree nodes and fewer high degree nodes, compared to those from the age-based strategy. Similar is observed between the snapshot network before and after the second vaccination. After that the degree distribution does not change much with vaccination, since the network is already very sparse.

**Figure 21:**
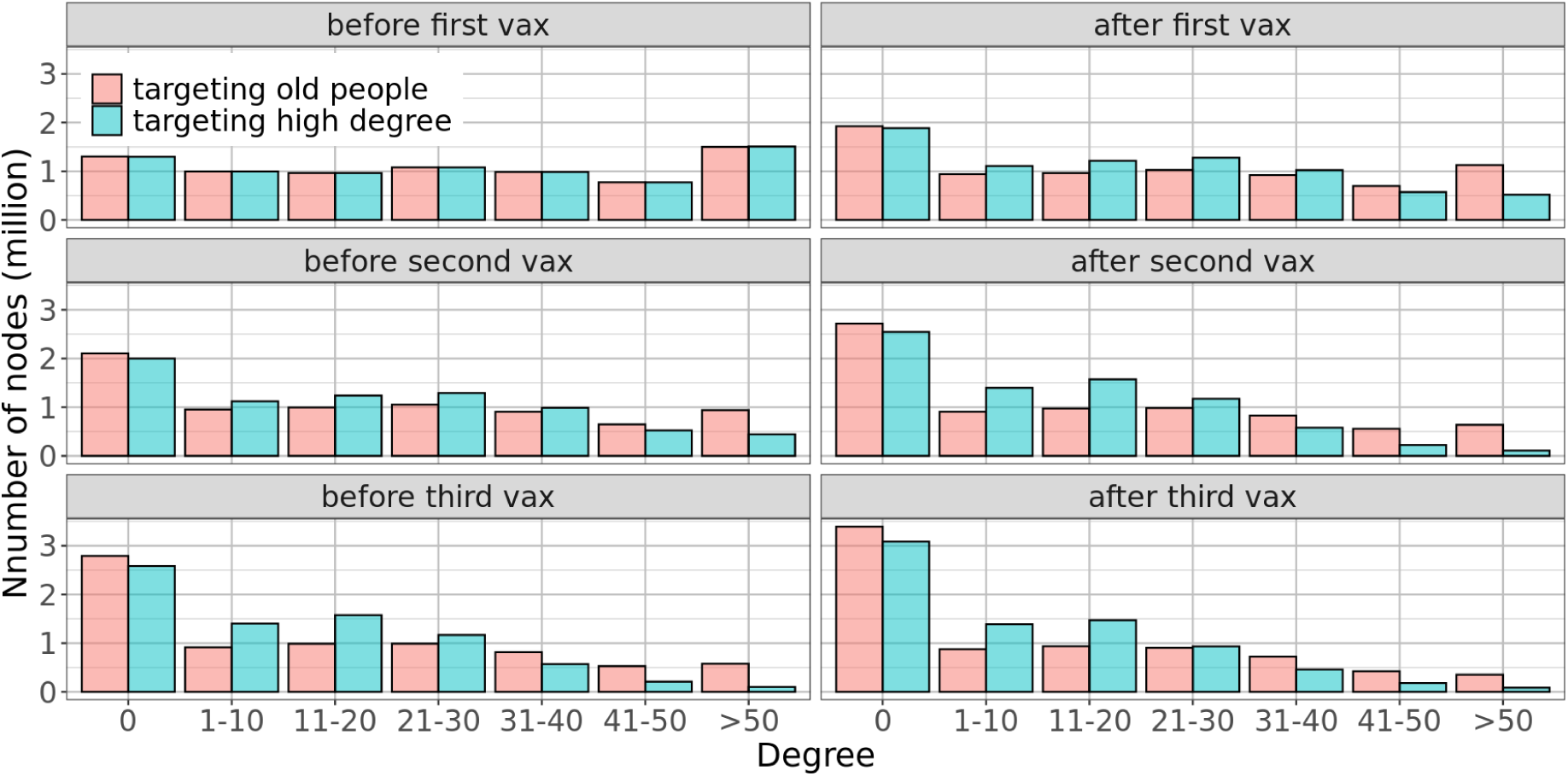
As nodes are infected or vaccinated, we remove edges incident on them from the contact network, to form sparser and sparser *snapshot* networks. We consider such snapshots under age-based and degree-based strategies, assuming no relaxation of NPIs and 100% distribution. After the first round of vaccination, the degree-based strategy reduces degrees more than the age-based strategy: we observe more low degree nodes and fewer high degree nodes in the snapshot networks from the degree-based strategy.

The connection between spectral properties and epidemic thresholds provides another insight into the efficacy of a degree-based strategy. As mentioned in Section B, it has been shown for SIS/SIR models, via different approaches (including different mean field approximations), that if the first eigenvalue (referred to as the *spectral radius*) *λ*_1_(*G*) of the network is below a certain threshold, the epidemic is not very “large” [27, 45, 52]. Figure 19a shows the first eigenvalue *λ*_1_(*G*) of the network at different points in time as interventions are applied. It is very significant to note that *λ*_1_(*G*) shows a substantial drop when the degree intervention is applied. Thus, the intuition behind the efficacy of such methods is clear – high degree nodes protect not only themselves, but also confer a higher level of indirect protection on their neighbors as they interact with many individuals. The fact that real-world social networks have sufficient nodes of high degree ensure that such heuristics are effective. It is important to note, however, that just the presence of high degree nodes themselves does not ensure that degree-based heuristics work.

### A dynamical property: Vulnerability and its relationship to degree

Given a generalized SEIR system *G*(*V, E*) along with a random initial configuration and a node *v ∈ V*; and an integer *t ≥* 1, we define *t*-Vul(*v*) as the probability that *v* gets infected *at* time *t*. Vulnerability of *v* by time *t* (*t*-TotVul(*v*)) is the probability that *v* is gets infected *by* time *t*. Informally speaking, *t*-Vul(*v*)and *t*-TotVul(*v*)estimate the chance of a node getting infected when a random set of nodes are infected to begin with. Vulnerability in essence captures various ways the infection can reach a given node. Degree-based interventions effectively reduce the vulnerability of several nodes at once. Interestingly, nodes of high degree are also highly vulnerable, and this can be empirically observed by noting the plots in Figures 22 and 23. Vulnerability computations are computationally challenging (`#P-hard`) and our simulations provide good estimates of the same.

**Figure 22:**
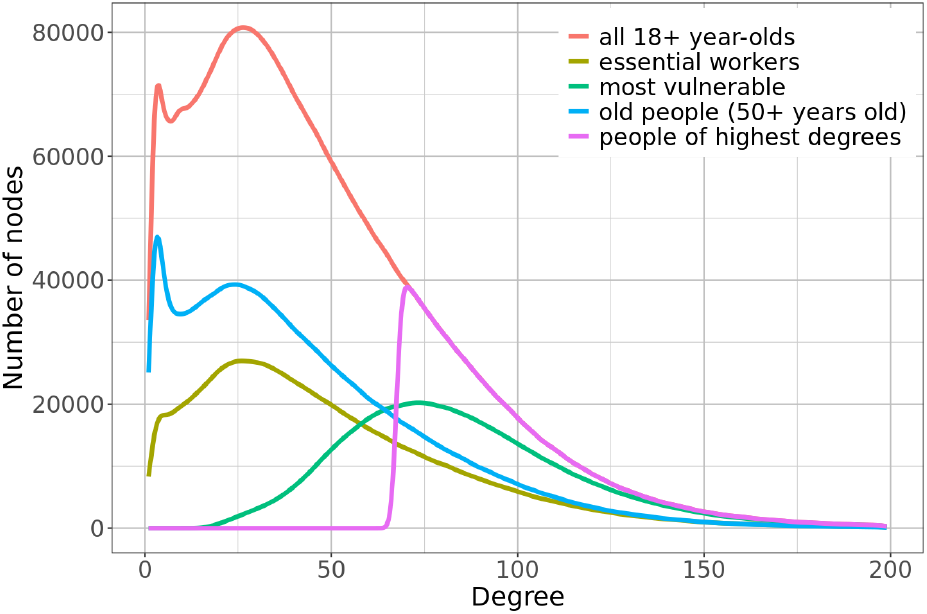
Degree distribution among nodes that are prioritized differently. Essential workers and older people seem to have lower degrees than high degree people and highly vulnerable people.

**Figure 23:**
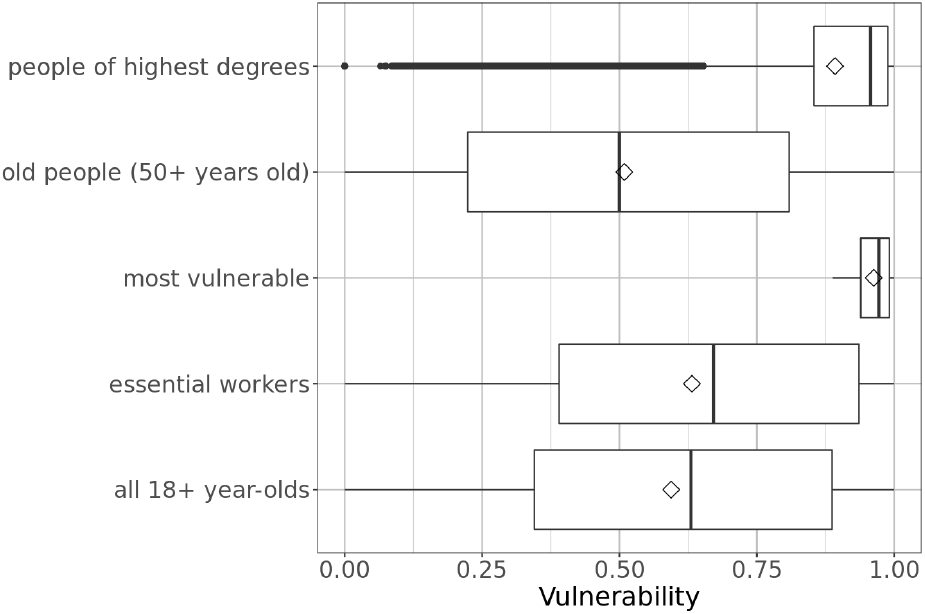
Vulnerability distribution of nodes with different priorities. People of highest degrees have high vulnerability, close to that of the most vulnerable people.

### How can we estimate degrees

A central question is: can individual node degrees be estimated? We believe that this is possible, and more so as a result of digital devices and apps that have recently been deployed for contact tracing. The current set of apps built for contact tracing keep track of the social interactions of an individual in a privacy-preserving way. These apps can easily be modified to count the number of such interactions. Furthermore, our results show that duplicate counts that might result are okay, as the degrees are likely to be skewed some. These apps can be modified to not only store the keys, but also the encrypted form of the time a person spends with the person that generates such a key.

Furthermore, a person does not need to reveal their degree to anyone but the app can simply provide this information for prioritization. The alerting and scheduling of vaccination appointments can be conducted through the app itself. Additionally, demographic information such as age, or other risk-factor information such as additional comorbidities or race and/or ethnicity, may also be used to further prioritize individuals within their degree-based allocation groups.

### How accurate can such measurements be

Let *V* be partitioned into groups *V*_1_*, . . . , V_r_*, and suppose *p_i_* is the probability that a node in *V_i_* has the app. Think of *V_i_* as an age group. Survey results show that younger individuals have a higher propensity to use digital device as well as download the app. We assume each node *v ∈ V_i_* decides independently with probability *p_i_* to install the app; if *v* chooses to install the app, we say that it is “sampled”. Let *H* denote the sampled graph, which is induced by the nodes (node induced subgraph) which have the app, and let *V* (*H*) denote the set of nodes in *H*. For a node *v*, let *N_G_*(*v*) denote the set of its neighbors in *G*. Similarly, let *N_H_* (*v*) = *{u*: *u ∈ N_G_*(*v*) and *u ∈ V* (*H*)*}* denote the set of neighbors of *v* in *H*. Let *d_G_*(*v*) = *|N_G_*(*v*)*|* denote its degree in *G*, and let *d_G_*(*v, i*) denote the number of neighbors it has in group 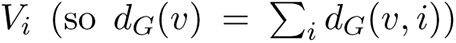. If *v* is sampled, let *d_H_* (*v*) = *|N_H_* (*v*)*|* denote its degree in *H*. Let *X*(*u*) ∈ {0, 1} be a random variable which is 1 with probability *p_i_*, if *u ∈ V_i_*, and let 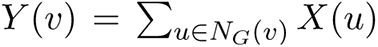. Observe that 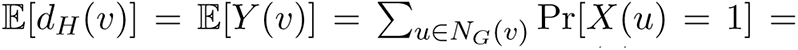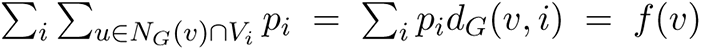. By applying a Chernoff bound to *Y* (*v*), we have 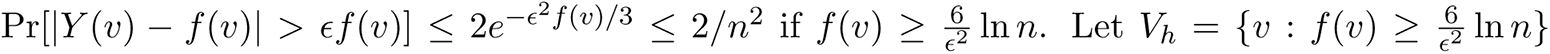 Therefore, with probability at least 1 *−* 2*/n*, every node 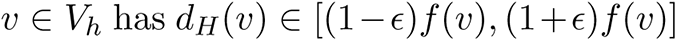. Let *Q* ⊆ *V* denote the subset of nodes in the top quartile with respect to *d_G_*(*v*). Then, with high probability, for every node *v ∈ V_h_ ∩ Q*, we have *d_H_* (*v*) *∼ f* (*v*). Therefore, if the sets *V_h_* and *Q* have high overlap, high degree in *H* corresponds to high degree in *G*. The expected number of nodes in *V_h_ ∩ Q* which get sampled is 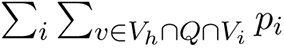

### Where is the tradeoff?

The phased approach is a balance between exposure/infection risk (policy equivalent of high social contact) and disease risk (age-based). Our proposal is a stronger version of the former, but seems to be significantly better across all outcomes. This is one of the counter-intuitive aspects of the proposed approach, since it does not take into account disease risk explicitly, and purely relies on structural measures of the network. While individualized policies aimed at minimizing mortality do exist (e.g., comorbidities) and are part of the phased approach, lack of technology thus far had made it difficult to ‘individualize’ policies targeted on minimizing transmission. Aggregate policies targeting essential workers and other high exposure categories (e.g., school children for influenza) while in a similar vein to our approach, may still have high variance in reducing transmission potential. The advantage of our approach is in leveraging the mechanistic and network based understanding of disease spread, and creating priority categories that cut across age-, risk- and other demographic characteristics. It is possible to construct pathological instances of networks in which vaccinating high degree nodes is not optimal. But as is well known in the field of network science, real world social networks are often characterized by power-law degree distributions and exhibit a hub-spoke structure. In such networks, targeting the hub nodes at individual level, ‘shatters’ the transmission network faster than aggregate policies. Especially such a strategy might be necessary in regions with very limited vaccine supplies and at higher risk of variant induced surges. We believe our work helps advance the case for better integration of social network and digital technologies in swifter public health response.

## F The Agent-Based Simulator

The EpiHiper agent-based simulator supports disease models which are comprised of *disease states*, *disease transmissions* (through contacts), and *disease progressions*. The disease models are specified independently of the people and their contact network over which the disease spreads. All individuals have the same infection processes and disease progression dynamics. The infection processes, however, factors in individual attributes such as susceptibility and infectivity which are generally dynamic. All input files to EpiHiper are provided in JSON format with the exception of the contact network which, due to its large size, uses either CSV or binary format.

**Disease Transmissions** are caused by contacts between a susceptible individual *P^s^* in state *X_i_* and an infectious individual *P^i^* in state *X_k_*. The susceptible individual *P^s^* will transition to an exposed state *X_j_* based on information specified by the transmission configuration *T_i,j,k_*, the contact *E*(*P^i^, P^s^*), and attributes from the individuals *P^s^* and *P^i^* such as the susceptibility *σ*(*P^s^*) of *P^s^* and the infectivity *ι*(*P ^i^*) of *P^i^*.

Under the assumption of independence of transitions across contacts for individual *P^s^* with one or more infectious individuals *P^i^*, the *propensity* of the state transition to the exposed state *X_j_* based on the transmission configuration *T_i,j,k_* and the single contact *E*(*P^i^, P^s^*) is defined as:

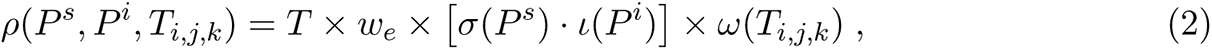

where *T* is the contact duration, *w_e_* is the edge weight, and *ω*(*T_i,j,k_*) is the transmission rate.

The *propensities* of all state transitions to the exposed state *X_j_* are added, and we use the Gillespie algorithm [30] to determine whether a transition occurs during the simulation interval, and, if it does, which contact to attribute.

### Disease Progression

covers the health state transitions within an individual 𝒫 that are independent of other people. For the EpiHiper model, a disease progression diagram describes all the possible health state transitions that take place within a person. The nodes of the diagram are the health states *X* = *{X_i_}* and directed edges of the form *e* = (*X_i_, X_j_*) with an assigned probability *p_e_* = prob(*X_i_, X_j_*) and a *dwell time distribution D_e_*. The sum of all probabilities of transitioning out of a given state *X_i_* must be either 1 or 0. Zero indicates a terminal state.

The **System State** at any point in time is given by the attributes of the individuals (nodes) and contacts (edges) of the network, the simulation time, and the value of user-defined variables.

The values *nodeTrait* and *edgeTrait* are user-defined attributes which may be used to govern interventions (described below). They do not influence the disease transmission or progression directly.

**Interventions** are external modifications of the state of the simulation where “external” means not governed by the disease model nor the contact network. An intervention comprises of a trigger, a target, and an action ensemble. The action ensemble, which is a set of instructions to apply to the dynamic state of the target (a collection of nodes and/or edges), is only applied when the trigger evaluates to 1 (or `true`). The trigger is a function of the system state and thus may depend on any of the above-mentioned attributes, including the *Person Trait Database*. The operations of an action ensemble are partitioned into the following three categories: (*i*) those triggered once per intervention (e.g., to update global variables), and which are thus independent of the intervention target, (*ii*) those that are applied to each element of the intervention target, and (*iii*) those applied to a sampled subset of the intervention target and optionally to the complement of the sampled subset. Moreover, sampling may be nested, thus permitting chained sub-sampling. Each individual action of the action ensemble is given a delay specifying the time (or offset) at which the action should execute, which permits flexible, fine-tuned scheduling within the simulation.

Before starting the simulation, the state of all individuals *P* must be initialized. In EpiHiper, initialization is simply a special case of an intervention where the trigger is omitted (and by convention is `true`).

## G The Disease Model Parameters

The within-host disease transmission model is shown in Figure 24. **Transmission** may occur when an individual in one of the states *Susceptible* or *RX Failure* comes in contact with one or more individuals in the states *Presymptomatic*, *Symptomatic*, or *Asymptomatic*. The individual transmissions are governed by the parameters in Table 5. **Progression** from one disease state to the next is governed by the parameters in Table 6

**Figure 24:**
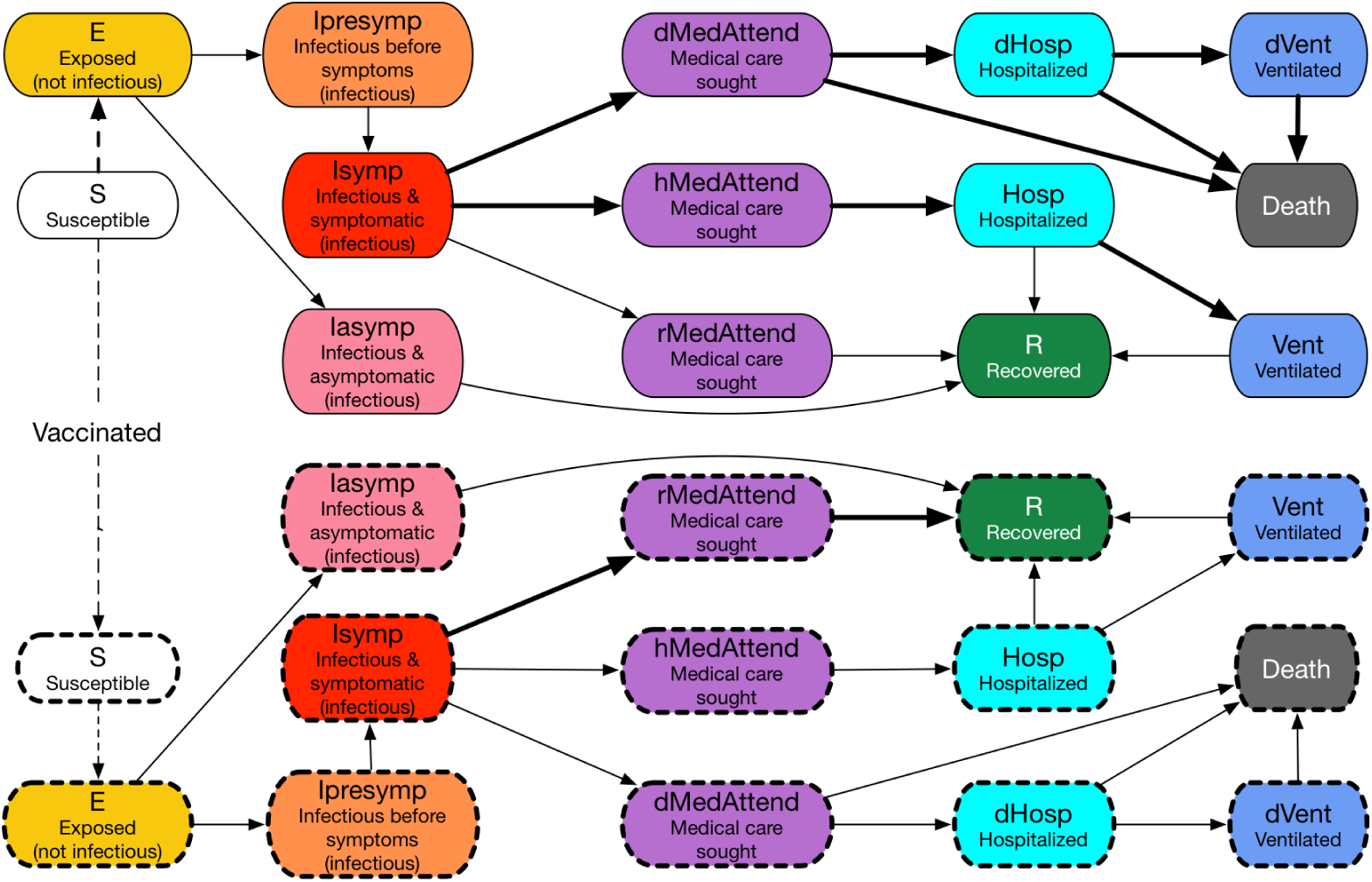
The COVID-19 disease model with both unvaccinated and vaccinated states. This disease progression model is represented as a probabilistic timed transition system (PTTS): the state transitions are probabilistic, and, in many cases, are timed, i.e., transitions after a given time period. An individual starts from the upper S (Susceptible) state. If an individual receives a vaccine the individual enters a new susceptible state represented by a dotted box. The dashed lines represent state transitions triggered by either interactions with infectious individuals or vaccination. The solid lines represent probabilistic timed state transitions. The shapes with a solid border represent states of an unvaccinated individual; those with a dashed border represent states of a vaccinated individual. The thicker lines represent larger probabilites. Therefore, if vaccinated, an individual has a smaller probability of getting infected (protection against infection), and even if infected the individual has smaller probability of being hospitalized or needing ventilation or death (protection against severe illness).

**Table 5:**
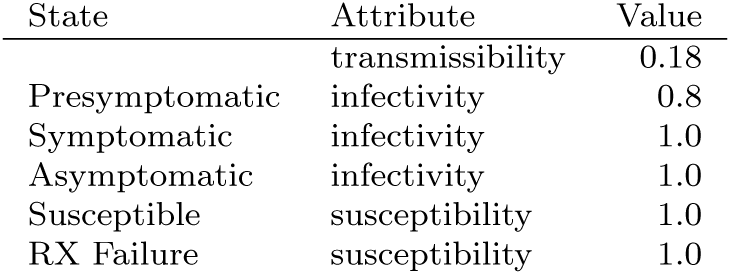
Disease transmission parameters

**Table 6:** Disease progression parameters as given by the CDC document [15]. One value per line applies to all age groups. Abbreviations: prob: probability, dt: dwell time, Attd: attended, Hosp: hospitalized, Vent: ventilated, (D): resulting in death, and (H): resulting in hospitalization

| Progression | Attribute | Age |  |  |  |  |
| --- | --- | --- | --- | --- | --- | --- |
|  |  | 0-4 | 5-17 | 18-49 | 50-64 | 65+ |
| Exposed - Asympt | prob |  |  | 0.35 |  |  |
| Exposed - Asympt | dt-mean |  |  | 5 |  |  |
| Exposed - Asympt | dt-std dev |  |  | 1 |  |  |
| Asympt - Recovered | prob |  |  | 1 |  |  |
| Asympt - Recovered | dt-mean |  |  | 5 |  |  |
| Asympt - Recovered | dt-std dev |  |  | 1 |  |  |
| Exposed - Presympt | prob |  |  | 0.65 |  |  |
| Exposed - Presympt | dt-fixed |  |  | 1 |  |  |
| Presympt - Sympt | prob |  |  | 0.65 |  |  |
| Presympt - Sympt | dt-fixed |  |  | 1 |  |  |
| Sympt - Attd | prob | 0.9594 | 0.9894 | 0.9594 | 0.912 | 0.788 |
| Sympt - Attd | dt-discrete | 1:0.175, 2:0.175, 3:0.1, 4:0.1, 5:0.1, 6:0.1, 7:0.1, 8:0.05, 9:0.05, 10:0.05 |  |  |  |  |
| Attd - Recovered | prob |  |  | 1 |  |  |
| Attd - Recovered | dt-mean |  |  | 5 |  |  |
| Attd - Recovered | dt-std dev |  |  | 1 |  |  |
| Sympt - Attd(D) | prob | 0.0006 | 0.0006 | 0.0006 | 0.003 | 0.017 |
| Sympt - Attd(D) | dt-fixed |  |  | 2 |  |  |
| Attd(D) - Hosp(D) | prob |  |  | 0.95 |  |  |
| Attd(D) - Hosp(D) | dt-fixed |  |  | 2 |  |  |
| Hosp(D) - Vent(D) | prob | 0.06 | 0.06 | 0.06 | 0.15 | 0.225 |
| Hosp(D) - Vent(D) | dt-fixed |  |  | 2 |  |  |
| Vent(D) - Death | prob |  |  | 1 |  |  |
| Vent(D) - Death | dt-fixed |  |  | 4 |  |  |
| Hosp(D) - Death | prob | 0.94 | 0.94 | 0.94 | 0.85 | 0.775 |
| Hosp(D) - Death | dt-fixed |  |  | 6 |  |  |
| Attd(D) - Death | prob |  |  | 0.05 |  |  |
| Attd(D) - Death | dt-fixed |  |  | 8 |  |  |
| Sympt - Attd(H) | prob | 0.04 | 0.01 | 0.04 | 0.085 | 0.195 |
| Sympt - Attd(H) | dt-fixed |  |  | 1 |  |  |
| Attd(H) - Hosp | prob |  |  | 1 |  |  |
| Attd(H) - Hosp | dt-mean | 5 | 5 | 5 | 5.3 | 4.2 |
| Attd(H) - Hosp | dt-std dev | 4.6 | 4.6 | 4.6 | 5.2 | 5.2 |
| Hosp - Recovered | prob, |  |  | 0.2 |  |  |
| Hosp - Recovered | dt-mean | 3.1 | 3.1 | 3.1 | 7.8 | 6.5 |
| Hosp - Recovered | dt-std dev | 3.7 | 3.7 | 3.7 | 6.3 | 4.9 |
| Hosp - Vent | prob | 0.06 | 0.06 | 0.06 | 0.15 | 0.225 |
| Hosp - Vent | dt-mean |  |  | 1 |  |  |
| Hosp - Vent | dt-std dev |  |  | 0.2 |  |  |
| Vent - Recovered | prob |  |  | 1 |  |  |
| Vent - Recovered | dt-mean | 2.1 | 2.1 | 2.1 | 6.8 | 5.5 |
| Vent - Recovered | dt-std dev | 3.7 | 3.7 | 3.7 | 6.3 | 4.9 |

## H Related Work

There has been a lot of work on analyzing interventions to control epidemics, and this falls into two broad categories. The first involves using a system of coupled differential equations to represent the dynamics, e.g., [42, 43, 55, 69, 72]. Even though closed-form solutions are not available even for simple models, when the system is not very large, it can be solved by brute-force local search methods, e.g., [43]. For some types of models, greedy strategies have been used [68, 72]. The second class of methods is network or agent based, of the form we study here, e.g., [23, 29, 31, 38, 40].

Analyzing interventions to minimize the expected outbreak size (or to optimize other epidemic outcomes) in network models is much harder, and is generally open to problems. Prior work has generally attempted to solve these problems by either simplifying the network (e.g., assuming random graph models), or simplifying the disease model. The simplest setting is that of transmission probability of 1 (modeling a highly contagious disease), with a fixed source. Even this setting is challenging, and work by [24, 32] designs bicriteria approximation algorithms for this problem.

A variation of this setting is when the source is chosen randomly, and, in this case, the problem of minimizing the number of infections corresponds to deleting a subset of nodes such that the sum of squares of the component sizes in the residual network is minimized. A minor modification of the results of [6, 36] gives approximation algorithms for this objective. We note that [58] uses a stochastic optimization approach for minimizing the expected number of infections. While their worst case approximation factor can be quite large, their empirical performance is quite good. The work of [3, 10] on the robustness of networks can be viewed as interventions to reduce the spread of an outbreak.

It is well understood that the network structure has a significant impact on the dynamics of epidemic spread. This has motivated a lot of research on modifying network properties to control epidemic spread. One of the most studied properties is degree, and in many network models, as well as in a broad class of real world networks, it has been found that removing the highest degree nodes (equivalently, vaccinating high degree nodes) turns out to be very effective [3,10,19,22,51,73]. Cohen et al [19] show that a simple decentralized strategy of “acquaintance immunization” has the effect of selecting high degree nodes. Another set of properties that has been studied extensively are spectral properties, namely the eigenvalues and eigenvectors associated with the adjacency matrix of the graph and its Laplacian. It has been shown using multiple approaches [27,45,52] that epidemic spread exhibits a threshold behavior—if the spectral radius (the largest absolute value of an eigenvalue) is below a certain threshold, the disease dies out. This has motivated a considerable amount of work on reducing the spectral radius to control the outbreak [46, 53, 54, 56, 62]. *In general, the theoretical studies do not apply to temporal vaccine allocation problems – in such cases the network is constantly changing as the epidemic spreads and vaccines are distributed in time*.

In the context of COVID-19, where we have multiple approved vaccine candidates, the role of vaccine efficacy, especially whether it reduces susceptibility to disease or transmission becomes important [37]. A recent study by Bubar et al. [12] identified that under different underlying assumptions, vaccine prioritization policies vary from 20-49 years to adults over 60 years old. They also note that prioritizing seronegative individuals could improve the marginal impact of a given policy. A similar study at a global scale using different supply assumptions was reported in [34]. See [1, 9, 26, 34, 41, 55, 59] for other recent papers on this topic. Multiple studies have also identified the tradeoffs based on the underlying policy objectives [13, 41] using compartmental models. The current allocation policy in the US at the federal level is centered around the framework developed by the National Academies of Sciences, Engineering, and Medicine (NASEM) [48].

Very few papers have studied vaccine allocation problems when there is a vaccine schedule (temporal vaccine allocation). Furthermore, they do not study how robust such methods are against uncertainty in estimating the structural properties; this is a crucial contribution of the present paper. Nevertheless, these results do suggest the potential value of such methods.

### Digital apps to estimate network properties

Digital contact tracing apps have recently been deployed in several countries [2, 5, 17, 25, 35] with mixed success. Reasons for the range of outcomes include: (*i*) low penetration levels, (*ii*) compliance, and (*iii*) accuracy of the apps in discovering neighbors accurately. Our allocation method is based on exploiting simple network properties that can be estimated using digital devices. We use two measures here: (*i*) degree and (*ii*) weighted degree. Digital contact tracing apps can potentially measure both of these quantities quite accurately.

## Footnotes

^1^See https://nssac.bii.virginia.edu/covid-19/dashboard/ for latest figures

^2^https://www.fda.gov/news-events/press-announcements/coronavirus-covid-19-update-fda-takes-action-help-facilitate-timely-development-safe-effective-covid

^3^https://landscan.ornl.gov/.

^4^http://www.openstreetmap.org

^5^https://sedac.ciesin.columbia.edu/data/collection/gpw-v4

^6^Note that though edge *e* is represented as a tuple (*u, v*), it actually denotes the set *{u, v}*, as is common in graph theory.

